# Sensitivity of Estimated Tacrolimus Population Pharmacokinetic Profile to Inaccurate Assumptions about Dose Timing and Absorption: An Investigation in Real-World and Simulated Data

**DOI:** 10.1101/2021.01.15.21249900

**Authors:** Michael L. Williams, Hannah L. Weeks, Cole Beck, Leena Choi

## Abstract

A population pharmacokinetic (PK) study with 363 subjects was performed using real-world data extracted from electronic heath records (EHRs) to estimate the tacrolimus population PK profile. As population PK studies for oral medications performed using EHR data often assume a regular dosing schedule as prescribed without incorporating exact dosing time, we assessed the sensitivity of the PK parameter estimates to assumptions about dose timing using last-dose times extracted by our own natural language processing system, *medExtractR*. We also investigated the sensitivity of estimations to absorption rate constants that are often fixed at a published value in tacrolimus population PK analysis. There was no appreciable difference in parameters estimates with *vs*. without last-dose time incorporated in the data and our sensitivity analysis revealed little difference between parameters estimated assuming a range of absorption rate constants. We also conducted simulation studies to investigate how drug PK profiles and experimental designs such as concentration measurements affects sensitivity to incorrect assumptions about dose timing and absorption rates. Our findings suggest that drugs with a slower elimination rate (or a longer half-life) are less sensitive to dose timing errors and that experimental designs which only allow for trough blood concentrations are usually insensitive to deviation in absorption rate.

## Introduction

Tacrolimus is an immunosuppressive calcineurin inhibitor medication widely used to prevent rejection following organ transplantation. Tacrolimus has a narrow therapeutic window, which makes it imperative for drug concentration levels to be appropriately maintained. Changes in dose, concomitant medications, or co-morbidities can yield less efficacy (i.e., an increased probability of allograft rejection)^1^ or more toxicity (e.g., nephrotoxicity, neurotoxicity, diabetogenicity).^2,3^ The challenge of maintaining this therapeutic range is exacerbated by individual variability in tacrolimus disposition. Several patient characteristics affecting clearance have been reported: examples include liver function, time since transplantation,^4^ and age.^5^ In particular, the effect of a single nucleotide polymorphism (SNP), rs776746, on the *CYP3A5* gene coding the major metabolizing enzyme for tacrolimus has been reported from multiple studies, including our own.^6^ Individuals carrying *CYP3A5*3*, a loss-of-function allele, have reduced clearance and require significantly smaller tacrolimus doses to maintain the same tacrolimus concentration level.^7–11^ This SNP alone was estimated to be associated with 39% of the variability in dose requirement between subjects.^6^

Population pharmacokinetic (PK) studies have traditionally relied on observational studies often prospectively performed during routine care. While such studies are not as costly or restrictive as randomized controlled trials, they still impose costs associated with data collection and restrictions on participant enrollment.^12^ Postmarketing drug studies have been increasingly performed using real-world data sources such as electronic health records (EHRs), which are particularly valuable as a source of longitudinal clinically relevant data. The adoption of EHRs has made large-scale retrospective studies feasible with rapid cohort generation from patients enrolled in routine care, leading to increased interest in using EHRs as real-word data.

We recently developed a natural language processing (NLP) system, *medExtractR*,^13^ to extract medication information from free-text clinical notes as part of a system to enable the use of EHRs in retrospective studies of drugs.^14,15^ The system, once finalized, should relieve the primary burden in data generation and manual extraction of medication data. In addition to drug dosing information, *medExtractR* is designed to extract explicit last dosing times (timing of the dose prior to a recorded blood concentration) if present in the notes. Lacking such data, tacrolimus population PK studies often assume dosing time at a regular dosing schedule as prescribed – e.g., b.i.d., or once every twelve hours for immediate-release formulation. We refer this to as the assumed dosing schedule, and any deviation of this assumed schedule from the actual dosing time may result in different estimates of the PK parameters. To the best of our knowledge, no studies have investigated the sensitivity of tacrolimus PK parameter estimates to deviations from the assumed dosing schedule.

Due to the narrow therapeutic window with high variability in tacrolimus disposition, tacrolimus is one of the drugs that requires therapeutic drug monitoring (TDM).^16^ Tacrolimus blood concentrations are routinely checked as part of standard of care, making retrospective studies with EHR data more appealing with clinically generated drug level data. Several tacrolimus population PK studies have been conducted using such data.^17–19^ As tacrolimus population PK studies are performed retrospectively using trough drug concentrations, they often assume the absorption rate constant, *k*_*a*_, at a fixed value in the PK models. Although some studies may perform a sensitivity analysis, the effect of this assumption has not been thoroughly investigated. We investigate this assumption by comparing models fit at varying assumed *k*_*a*_ levels. The relevance of these concerns to future work is modified by the increasing use of an extended-release formulation of tacrolimus^20^ which has a different PK profile to the immediate-release formulation and therefore may be more or less sensitive to the assumptions we are investigating. While the investigation of this new formulation is out of scope of this study, our findings may give insights into how assumptions and experimental design might impact PK parameter estimation for extended-release formulations as well.

The goals of this study were fourfold: (1) to identify the important factors affecting tacrolimus PK profile through population PK analysis using real-world data extracted from EHRs; (2) to investigate the effects of last dosing time on the tacrolimus PK parameter estimates using the data extracted from the EHRs; (3) to investigate the sensitivity of assumption to the absorption rate constant, *k*_*a*_, at a fixed value in a model on the other PK parameter estimates; and (4) to investigate the effects of last dosing time and assumption of fixed *k*_*a*_ on PK parameter estimates using simulation studies.

## Methods

### Study Design and Data Source

This study was approved by the Vanderbilt Institutional Review Board. We used data obtained from the previous study, which was described in detail in Birdwell *et al*.^6^ Briefly, medical records for individuals who received a kidney transplant were identified in the Vanderbilt Synthetic Derivative, a deidentified version of Vanderbilt’s EHR. These patients were matched to blood samples in BioVU, a biorepository of DNA collected during routine clinical testing. Patients having at least three measured tacrolimus blood concentrations, evidence of tacrolimus treatment in their medical records, and available tacrolimus dosing data were used for genotyping. These 527 subjects were further reduced either due to genotyping quality control or the presence of ancestry availability markers in BioVU to yield the final sample of 399 subjects. While the original study collected extensive genotype information, we restrict our interest to the genetic effect of mutations in *CYP3A5*. Genotyping was done to detect *CYP3A5*1*3* or *CYP3A5*3*3* mutations and, if none were detected, *CYP3A5*1*1* was assumed to be the patient genotype. Note that no tacrolimus extended-release formulation but only immediate-release formulation was prescribed during the study period. We draw our cohort from that sample, making use of tacrolimus doses, drug concentration levels, demographics, laboratory, and genotype data. Additionally, we separately extracted the last dosing times from the same clinical notes that yielded the original dataset.

### Extraction of Last-Dose Times

Times of last dose were extracted using *medExtractR* (see Weeks *et al*.^13^ for details). **Figure 1** outlines the algorithm to process these extractions. Extracted last dose times appeared in various formats, for example using am or pm (e.g., “10 am”, “9:30 pm”), military time (e.g., 2200, 20:30), or using a modifying word or phrase (e.g., “8 last night”, “yesterday morning at 7”). All time expressions were initially converted into the same format of HH:MM:SS based on a 24 hour clock. For example, the phrase “8:30pm” would become “20:30:00”. Concentration measurements were generally assumed to be trough levels taken at a morning appointment. For this reason, PM last dose times were assumed to have occurred on the previous day and AM times were assumed to have occurred on the same day as the laboratory value. Conflicts could occur if different last dose times were extracted within the same note or on the same date. We observed a small percentage of conflicts (6.04%), about half of which (3.89%) were within 2 hours. Assuming concentration measurements were intended to be trough values, time differences within 2 hours were considered close enough to be equivalent with respect to impact on the PK parameter estimates and the earlier extracted time was kept. Cases where discrepancies still existed after removing differences within 2 hours (1.90%) were manually reviewed with a clinical expert to determine which last dose time was correct. Ambiguous cases where the correct last dose time could not be determined were treated as missing (0.90%).

**Figure 1.**
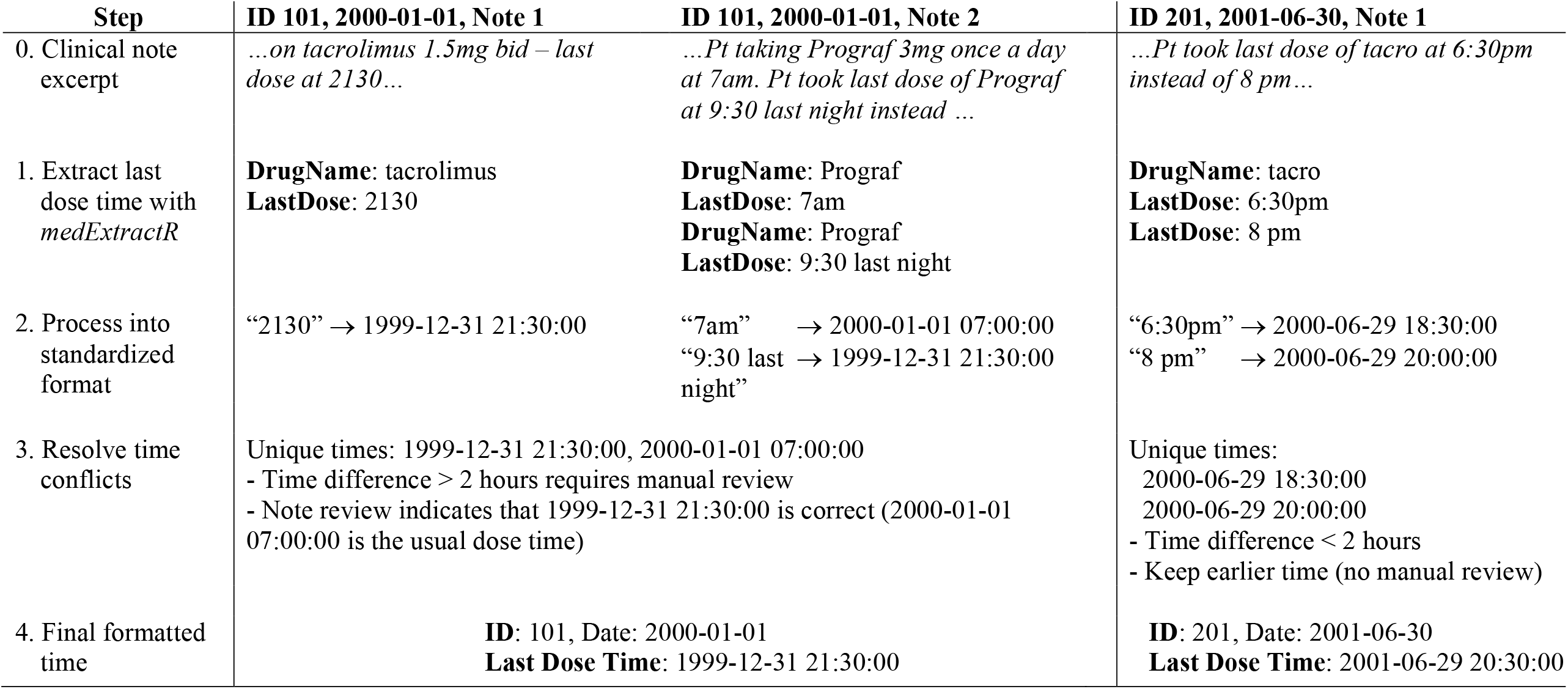
Last-Dose Time Extraction. Extraction and processing examples for time at which a subject took their last medication dose. In step 2, PM times are assumed to occur the day prior to the note date. AM times are assumed to occur on the same day at the note date. Final dataset includes one last dose time for each ID-date pair.

### Data Process

From the set of subjects (N = 399) described previously by Birdwell *et al*.,^6^ we refined our study cohort, called “entire cohort” (N = 363), as follows. We excluded drug concentration data measured before one month post-transplant or beyond three years post-transplant and then excluded subjects with fewer than 4 remaining concentrations. A subject with no known genotype for rs776746 (N = 1) was also excluded. We called this data as *the entire cohort without last-dose time* (which do not have NLP-extracted last-dose times). Next, last-dose times were extracted from the clinical notes for this same cohort to build a second dataset, called as *the entire cohort with last-dose time*. A reduced cohort was further defined based on the number of extracted last-dose times for each subject, excluding subjects having less than 4 concentration measurements with corresponding extracted last-dose times. This reduced cohort again has two datasets, called *the reduced cohort with last-dose time* and *the reduced cohort without last-dose time*. Thus, a total of four datasets, two for each cohort, were defined. Four PK datasets were built with an algorithm modified from a function (with an option for last-dose time) in an R package, *EHR*.^21^ A brief description of the PK data building method with and without last-dose time is in the next section below.

### PK Data Building

When last-dose time is not available within the clinical notes, a regular dosing interval is assumed as prescribed. Specifically, we assume that a dose of tacrolimus is taken 30 minutes (a reasonable timeframe confirmed by a clinician) after blood is drawn for the drug level checking. We further assume that the drug is taken every 12 hours following that initial dose, as all subjects took immediate-release formulation. This continues until the next concentration measurement, typically taken during morning clinic visits. The timing of this concentration measurement then determines the dose timing pattern until the next measurement is taken, and so on.

When last-dose time is available, the extracted time is used. For example, if a last-dose time is found matched with a concentration measurement, the preceding dose is set to be given at the time reflected in the clinical note. This dataset better represents the dosing times, among which the last-dose time is the most informative to estimate PK parameters. Note that not all extracted concentrations have an associated last-dose time available in the clinical notes. The datasets with and without last-dose time will be identical for the subset of concentration measurements missing last-dose time.

### Population PK Analysis

We performed population PK analysis of tacrolimus using a nonlinear mixed-effects model implemented by NONMEM®^22^ version VII with the first order conditional estimation method with interaction. A one-compartment PK model was chosen as the base model, assuming a combined additive and proportional residual error model and lognormal distribution for the random effects PK parameters. A model with random effects with unstructured covariance for all main PK parameters except for the absorption rate constant, *k*_*a*_, was assumed. As *k*_*a*_ cannot be reliably estimated without drug concentrations measured during the major absorption phase, it was fixed at the previously published value of 4.5.^23^

We considered the dataset of *the entire cohort with last-dose time* as the primary dataset used in the primary PK analysis to develop a population PK model. Covariate model building was performed using individual specific PK parameters estimated from the base model. Both graphical and statistical methods were considered with the following candidate covariates, which were chosen *a priori* based on previous research and biological plausibility: weight, age, sex, hemoglobin, albumin, race, and a *CYP3A5* SNP (rs776746). Model selection was performed based on the objective function values (−2 log likelihood). The difference in objective functions for models fit with and without a single covariate follows a χ^2^ distribution with one degree of freedom under the assumption of no covariate effect. The χ^2^ statistics of 3.84 with 1 degree of freedom corresponds to a p-value of 0.05. Thus, we considered an objective function value decrease of 3.84 to be significant model improvement. Variable selection was performed only in the primary analysis with the dataset of the entire cohort with last-dose time; selected variables were then used to build the same model from the remaining three datasets. The model was qualitatively assessed through visual examination using goodness-of-fit plots such as the observed vs. predicted concentrations, the conditional weighted residuals, and the visual predictive check that was performed using an R package, *vpc*.^24^

### Sensitivity Analysis

The *k*_*a*_ in our models was assumed to be 4.5 as previously reported.^23^ In order to assess whether our findings are sensitive to this selected value, we refit the model using the entire cohort at another published *k*_*a*_ value of 3.09.^10^ In addition, we refit the model with the published *k*_*a*_ value for the extended-release formulation, 0.375.^25^

### Simulation Study

In order to assess how different population PK characteristics and study design affect robustness to inaccurate last-dose times and absorption rate assumptions, we used two distinct PK profiles which we refer to as slow-elimination (SEL, elimination rate constant *k*_*e*_ = 0.02) and fast-elimination (FEL, *k*_*e*_ = 0.1). SEL approximates the PK profile of immediate-release tacrolimus as reported in Zuo *et al*.^10^ and FEL approximates that of immediate-release theophylline, as estimated from the well-studied and widely available theophylline dataset.^26,27^ We assume that SEL and FEL have a true underlying PK profile defined by a one-compartment, first order absorption/elimination rate constant model with combined additive and proportional residual error and random effects on clearance and volume of distribution.

True underlying PK parameter values defined for SEL and FEL are shown in **Table 1**. In addition, for each of SEL and FEL, two different levels of variability are defined by taking low or high variance components representing either a population with homogenous or heterogeneous PK profiles and low or high measurement error. This makes four major population profiles, with each PK profile having a low- and high-variance version. We term these SEL-HV, SEL-LV, FEL-HV, and FEL-LV. **Figure 2A** summarizes simulation scenarios along with the nomenclature and abbreviations used to describe the various simulation scenarios.

**Table 1.** PK Profiles for Simulation Study. True underlying parameters and dose (D) information are defined for two PK profiles with slow-(SEL) and fast-elimination (FEL). A total of four profiles, two scenarios of variability (low, high) for each of SEL and FEL, are created by taking either the low or high values for *ω*s and σs. The *k*_*e*_, *CL, V*, and *k*_*a*_ represent population PK parameters for elimination rate constant, clearance, volume of distribution, and absorption rate constant, respectively. The *ω*_*CL*_ and *ω*_*V*_ are the between-subject variance components for clearance and volume of distribution, presented as %CV. The *σ*_*prop*_ and *σ*_*add*_ represent the proportional and additive residual errors in the combined residual error model, presented as %CV and the standard deviation, respectively.

| Parameter | Slow-Elimination<br>(SEL) | Fast-Elimination<br>(FEL) |
| --- | --- | --- |
| $k_e$ (h<sup>-1</sup>) | 0.02 | 0.1 |
| $CL$ (L/h) | 22.7 | .2 |
| $V$ (L) | 1090 | 2 |
| $k_a$ (h<sup>-1</sup>) | 3.09 | 1 |
| $\omega_{CL}$ (%CV) | 5 or 30 | 5 or 30 |
| $\omega_V$ (%CV) | 5 or 30 | 5 or 30 |
| $\sigma_{prop}$ (%CV) | 5 or 30 | 5 or 30 |
| $\sigma_{add}$ (ng/ml) | 0.1 or 0.5 | 1 or 5 |
| D (mg) | 5 | 300 |

**Figure 2.**
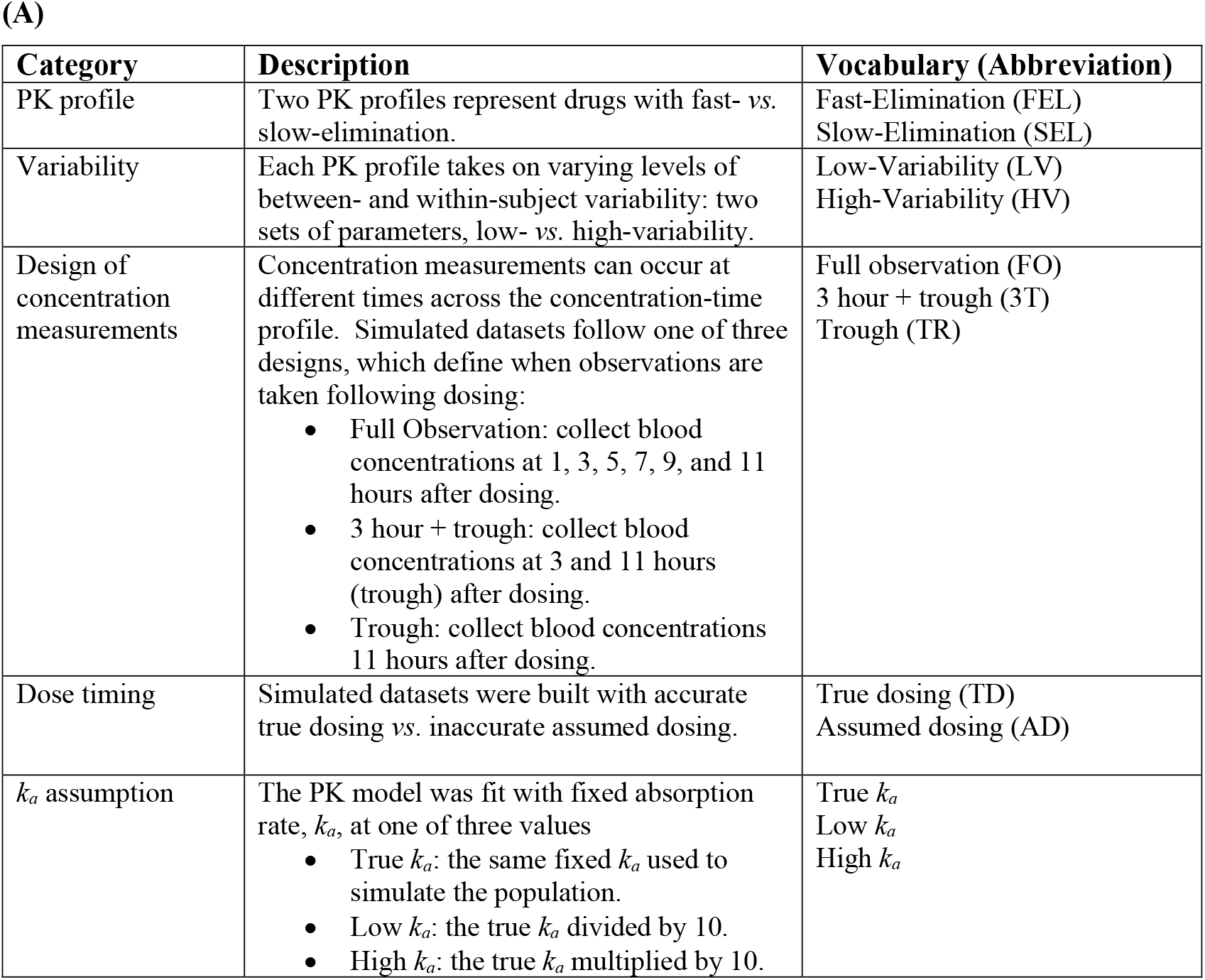
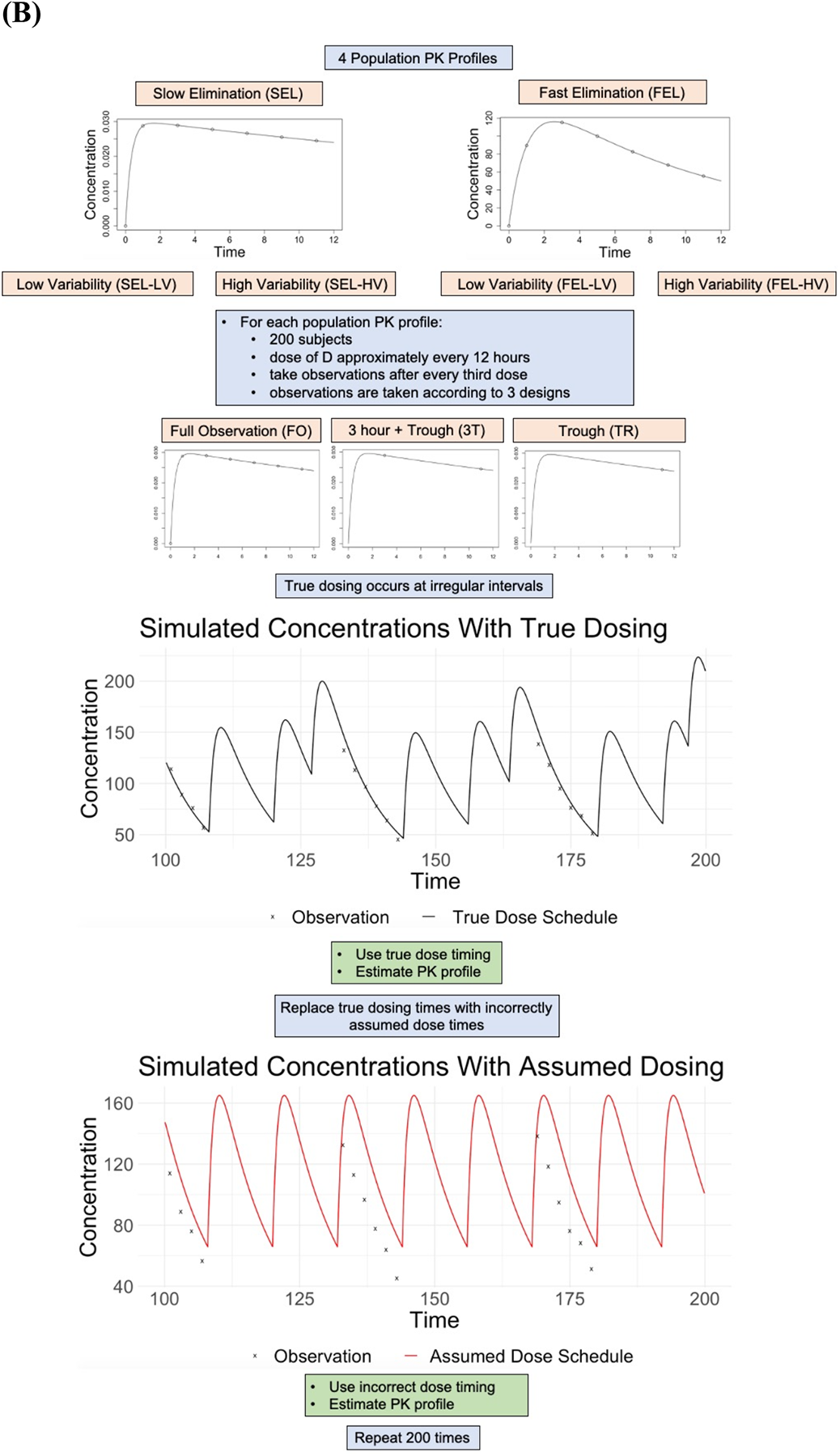
Outline of the Simulation Study to Investigate the Effect of Last-Dose Time Errors and Absorption Rate Assumption on PK Parameter Estimation. A description of scenarios along with nomenclature (A) and a diagram describing a simulation (B). Blood concentrations are simulated using various PK profiles with specific values defined in **Table 1**.

The simulation for investigating the impact of misspecified last-dose times on parameter estimates is outlined in **Figure 2B** and described as follows.

1. For each PK profile, 200 subjects are simulated as taking a dose (D) orally every 12 hours, called the “original every-12-hour dosing time”. A sufficient number of doses are given so that drug concentrations have reached steady state (i.e., 20 doses for SEL, 5 doses for FEL), then for the following 30 doses:
  a. True dosing (TD) time was generated by mimicking patients’ dose taking behavior as follows. Medication is taken every 12 hours but on every third dose dosing times are shifted by a random amount, *T*, which follows a normal distribution with mean -4 hours with standard deviation of 2 [i.e., *T* ∼ *Normal* (−4,2)]. These shifted dosing times define TD.
  b. Drug concentration measurements are simulated following these shifted dosing times (i.e., TD) according to three designs:
    i. Full observation (FO): Six observations measured at 1, 3, 5, 7, 9, and 11 hours following the original every-12-hour dosing time (i.e., before being shifted to TD).
    ii. 3-hour plus trough observations (3T): Two observations measured at 3 and 11 hours (which we consider the trough drug level) following the original every-12-hour dosing time.
    iii. Trough observation (TR): One observation measured at 11 hours following the original every-12-hour dosing time.
  c. Assumed dosing (AD) time is then generated by reversing shifted dose-times to the original every-12-hours dosing. This yields additional datasets with the same concentration measurements as above but incorrectly assumed dose times. This mimics PK datasets built from EHRs which usually do not have actual dose intake time information and hence must assume periodic dosing with some error.
2. The simulated data from each scenario is fit using a model with *k*_*a*_ fixed at its true value. The simulation for investigating the impact of incorrect specification of *k*_*a*_ proceeds similarly.
  1. For each PK profile, 200 subjects are simulated as taking a dose (D) orally every 12 hours. A sufficient number of doses are given so that drug concentrations have reached steady state (i.e., 20 doses for SEL, 5 doses for FEL), then for the following 30 doses blood concentrations are simulated according to the three previously described designs.
  2. Fit each simulated dataset with the same model assuming the following absorption rate constant, *k*_*a*_:
    a. *k*_*a*_ set to its true value (True *k*_*a*_).
    b. *k*_*a*_ set to one tenth of its true value (Low *k*_*a*_).
    c. *k*_*a*_ set to ten times its true value (High *k*_*a*_).

All simulations were performed with 200 repetitions for each scenario, and parameter estimates as well as the percent bias were evaluated. Data is simulated using R version 4.0.2 and models are fit using the stochastic approximation expectation-maximization estimation method implemented in Monolix® 2021.

## Results

### Population Characteristics for the Two Cohorts

The study population characteristics for the entire cohort and the reduced cohort are displayed in **Table 2**. Total number of concentrations for the entire cohort (N=363) and the reduced cohort (N=223) were 3258 and 2116, respectively. Each subject has a maximum of 10 tacrolimus blood concentration measurements. These observations constitute either the first 10 concentrations or every concentration if there are fewer than 10 measurements for each subject beyond the 1-month post-transplant period. Of these concentration measurements, 48% were accompanied by an extracted last-dose time for the entire cohort. The median tacrolimus dose across all subjects was 3 mg twice daily, and the median blood concentration was 7.5 ng/mL. For the reduced cohort, the percentage of concentration measurements associated with a last-dose time is increased to 73% as designed, while median tacrolimus dose and median blood concentration remain almost the same.

**Table 2.** Demographic and Clinical Characteristics. Population characteristics of the entire dataset (left) and the reduced dataset (right). Values are presented as count (proportion) for categorical variables and mean (SE) median [interquartile range] for continuous variable.

|  | Entire Cohort (N=363) | Reduced Cohort (N=223) |
| --- | --- | --- |
| Age (Year) | 45.9 (12.7)<br>47.0 [37.0, 55.0] | 47.2 (12.0)<br>48.0 [38.0, 55.0] |
| Sex = Female | 134 (0.37) | 76 (0.34) |
| Race |  |  |
| Caucasian American | 266 (0.73) | 164 (0.74) |
| African American | 83 (0.23) | 50 (0.22) |
| Hispanic | 4 (0.01) | 3 (0.01) |
| Asian | 5 (0.01) | 3 (0.01) |
| Other | 3 (0.01) | 3 (0.01) |
| Undisclosed | 2 (0.01) | 0 (0.00) |
| Weight (kg) | 82.9 (20.8)<br>81.2 [66.7, 95.3] | 83.0 (20.7)<br>80.0 [66.4, 94.9] |
| Hemoglobin (g/dL) | 12.5 (2.1)<br>12.5 [11.1, 14.0] | 12.6 (2.0)<br>12.5 [11.1, 14.0] |
| Albumin (g/dL) | 4.1 (0.4)<br>4.1 [3.9, 4.4] | 4.1 (0.4)<br>4.1 [3.8, 4.4] |
| Proportion of concentrations with last-dose time | 0.48 | 0.73 |
| Daily tacrolimus dose (mg/day) | 3.4 (1.8)<br>3 [2, 4] | 3.1 (1.7)<br>3 [2, 4] |
| Tacrolimus blood concentration (ng/mL) | 7.5 (3.1)<br>7.1 [5.5, 9.0] | 7.5 (3.0)<br>7.1 [5.5, 8.9] |
| Number of concentration levels per subject | 9.0 (1.8)<br>9.0 [10.0, 10.0] | 9.5 (1.2)<br>10.0 [10.0, 10.0] |
| Total number of concentrations | 3258 | 2116 |
| Total days of follow-up | 588 (295)<br>557 [361, 854] | 525 (268)<br>482 [324, 686] |

### Primary Population PK Analysis

**Table 3** presents all PK analyses results, from which we first describe the primary analysis using the entire cohort with last-dose time in this section. PK parameters such as clearance (*CL*, L/hr) and apparent volume of distribution (*V*, L) were first estimated from the base model without covariates. Note that we denote *CL/F* by *CL* and *V/F* by *V* for simplicity, where *F* represents relative bioavailability and is omitted elsewhere. The PK parameters varied substantially among subjects; the between-subject variation in coefficient of variation (CV) for *CL* and *V* in the base model are 52.8% and 65.5%, respectively.

**Table 3.**
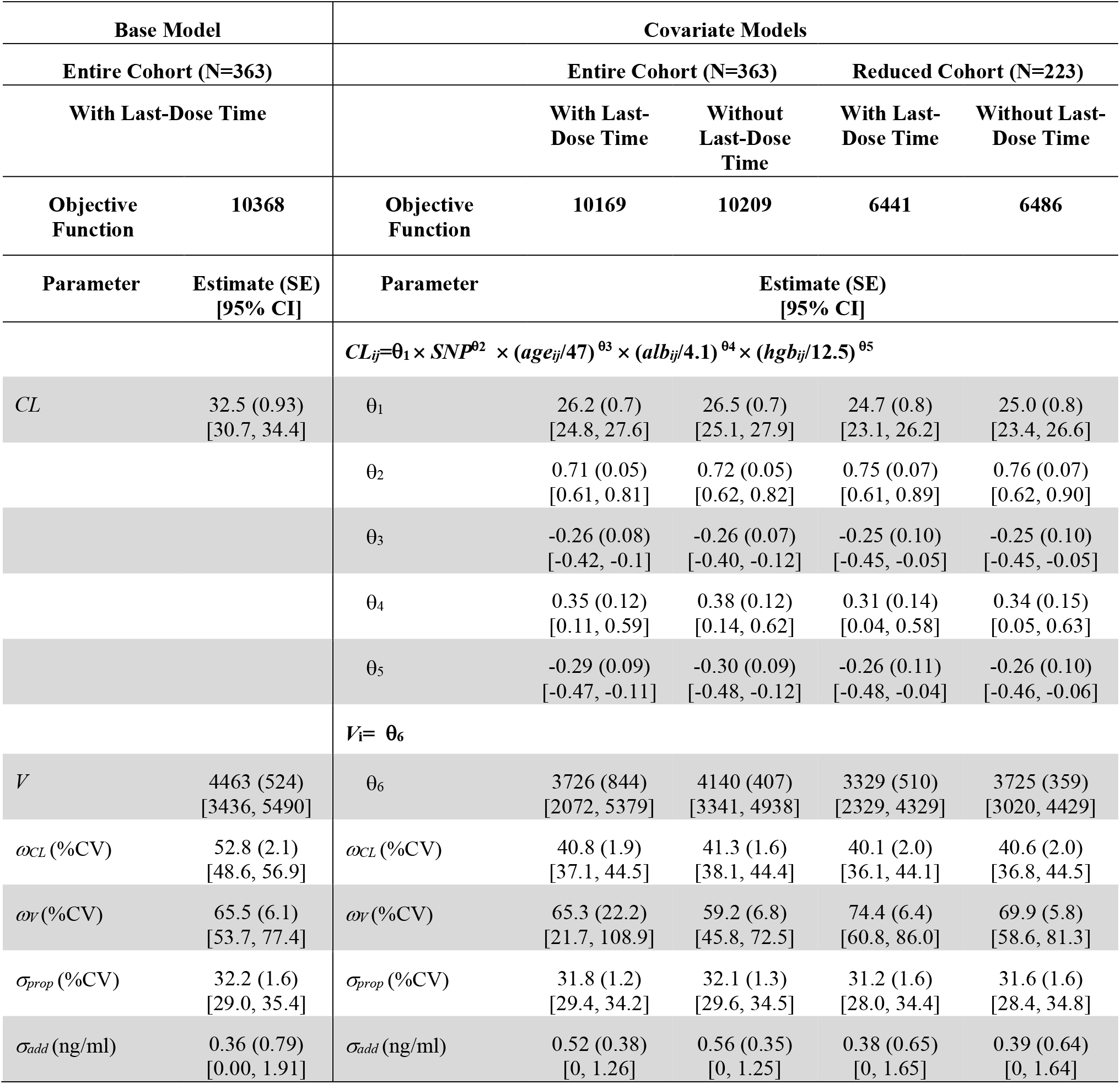
Estimated PK Parameters. PK parameter estimates for both the entire and reduced datasets either with or without last-dose time. Estimates are presented as mean (SE) [Wald 95% confidence interval]. *SNP* is coded continuously as 1, 2, 3 for 0, 1, and 2 mutations, respectively. The *age*_*ij*_, *alb*_*ij*_, and *hgb*_*ij*_ represent covariates values for subject *i* at time *j. CL* and *V* are population PK parameters for clearance and volume of distribution in the base model. The *ω*_*CL*_ and *ω*_*V*_ are the between-subject variance components for clearance and volume of distribution, respectively, presented as %CV. The *σ*_*prop*_ and *σ*_*add*_ represent the proportional and additive residual errors in the combined residual error model, presented as %CV and the standard deviation, respectively. The *θ*s denote model parameters as typically used in statistical models.

Inclusion of the SNP rs776746 alone decreased the objective function by 144 from 10368 for the base model; hence the SNP was always included in the following covariate model selection considering strong evidence for its causal effect on clearance in the literature as well as our own results. Inclusion of age, albumin level, and hemoglobin level all improved the model fit from the SNP only model – decreasing the objective function value by 18, 10, and 20, respectively. The final covariate model with these 3 additional covariates improved the model fit by 55 from the SNP only model. Other covariates did not yield significant improvement in the model fit. The final covariate model is presented as follows:

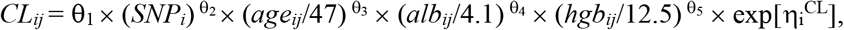

and

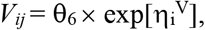

where *CL*_*ij*_ and *V*_*ij*_ are the subject-specific *CL* and *V* for subject *i* at time *j*. The *age*_*ij*_ is subject age in years, *alb*_*ij*_ is blood albumin level in g/dL, and *hgb*_*ij*_ is blood hemoglobin level in g/dL for subject *i* at time *j*, each of which is standardized by dividing by its population median. The genotype for SNP, rs776746, is continuously coded as 1, 2, 3 corresponding to 1 plus the number of *1 alleles in *CYP3A5*. The η_i_^CL^ and η_i_^V^ are random effects explaining between-subject variability for *CL* and *V*, which follow a bivariate normal distribution with mean zeros and covariance matrix with *ω*^*2*^_*CL*_ and *ω*^*2*^_*V*_ in its diagonal. The θs in the equations denote model parameters as typically used in statistical models.

The estimates of typical values of *CL* and *V* were 26.2 L/hr and 3726 L for a 47-year-old with no *\*1* allele in *CYP3A5*, albumin 4.1 g/dL, and hemoglobin 12.5 g/dL. Subjects with *\*1/*3* and *\*1/*1* increased clearance 1.6 (i.e., 2^0.71^ =1.6) and 2.2 (i.e., 3^0.71^ =2.2) times compared to clearance for those with *\*3/*3*, respectively. The between-subject variation in %CV for *CL* and *V* were 40.8% and 65.3%, respectively. Especially, the variability for *CL* was substantially reduced from the base model estimate of 52.8% as the inclusion of covariates explains a part of variability in clearance.

**Figure 3** shows model diagnostics. Overall, the goodness-of-fit plots present reasonable model fit although slight deviation from normality was noticed in the observed *vs*. predicted concentration plots. The weighted residuals show some deviation from normality, but most residuals fall within the range of -2 to 2. The visual predictive check also supports reasonable model fit in that the simulation-based model predicted 95% confidence intervals (CIs) well cover the corresponding observed values across the follow-up time.

**Figure 3.**
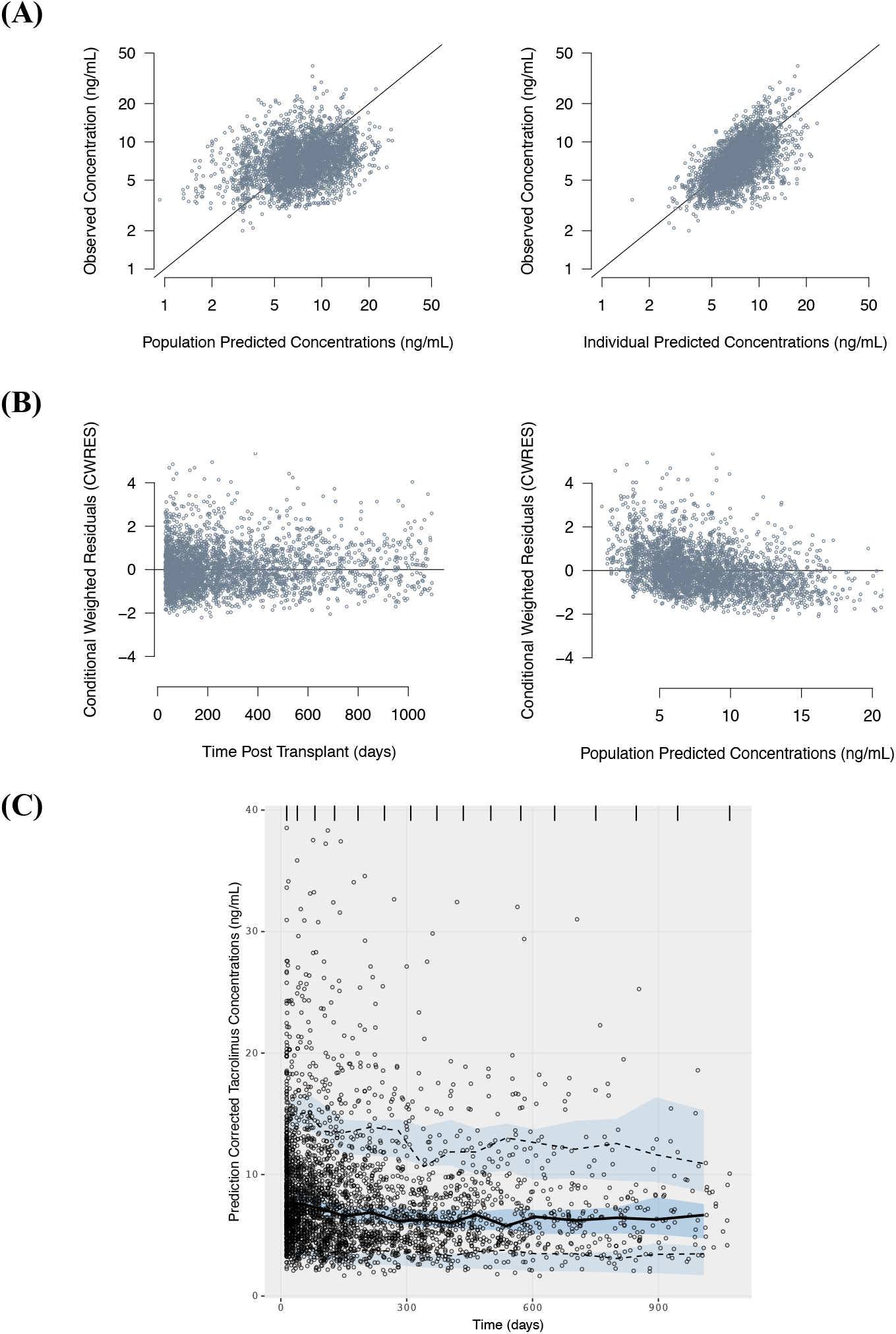
Model Diagnostics. Model diagnostic plots for the model fit to the entire cohort dataset with last-dose times. (A) Observed vs. predicted concentration plots on the log scale for both population (left) and individual (right) level predictions; (B) Conditional weighted residuals plots against time (left) and population predicted concentration (right); (C) the visual predictive check, where the solid line represents the median observed tacrolimus concentration while the dashed lines represent the 10th and 90th percentiles of the observed concentrations. Blue regions surrounding each line represent simulation-based model predicted 95% confidence intervals for the corresponding percentile.

### Last-Dose Time Inclusion

**Table 3** also presents parameter estimates in both the entire and reduced cohorts with and without last-dose times. There were little differences in parameter estimates with and without last-dose times within the same cohort. While the absolute difference in the estimates is greater between the cohorts given the last-dose time data status, the estimates across all four datasets are very similar and most 95% Wald CIs are overlapping. **Figure 4** shows the difference in extracted last-dose times and assumed last-dose times generated by a regular dosing assumption as prescribed. The median difference was -3 hours, suggesting actual doses were typically taken earlier than assumed.

**Figure 4.**
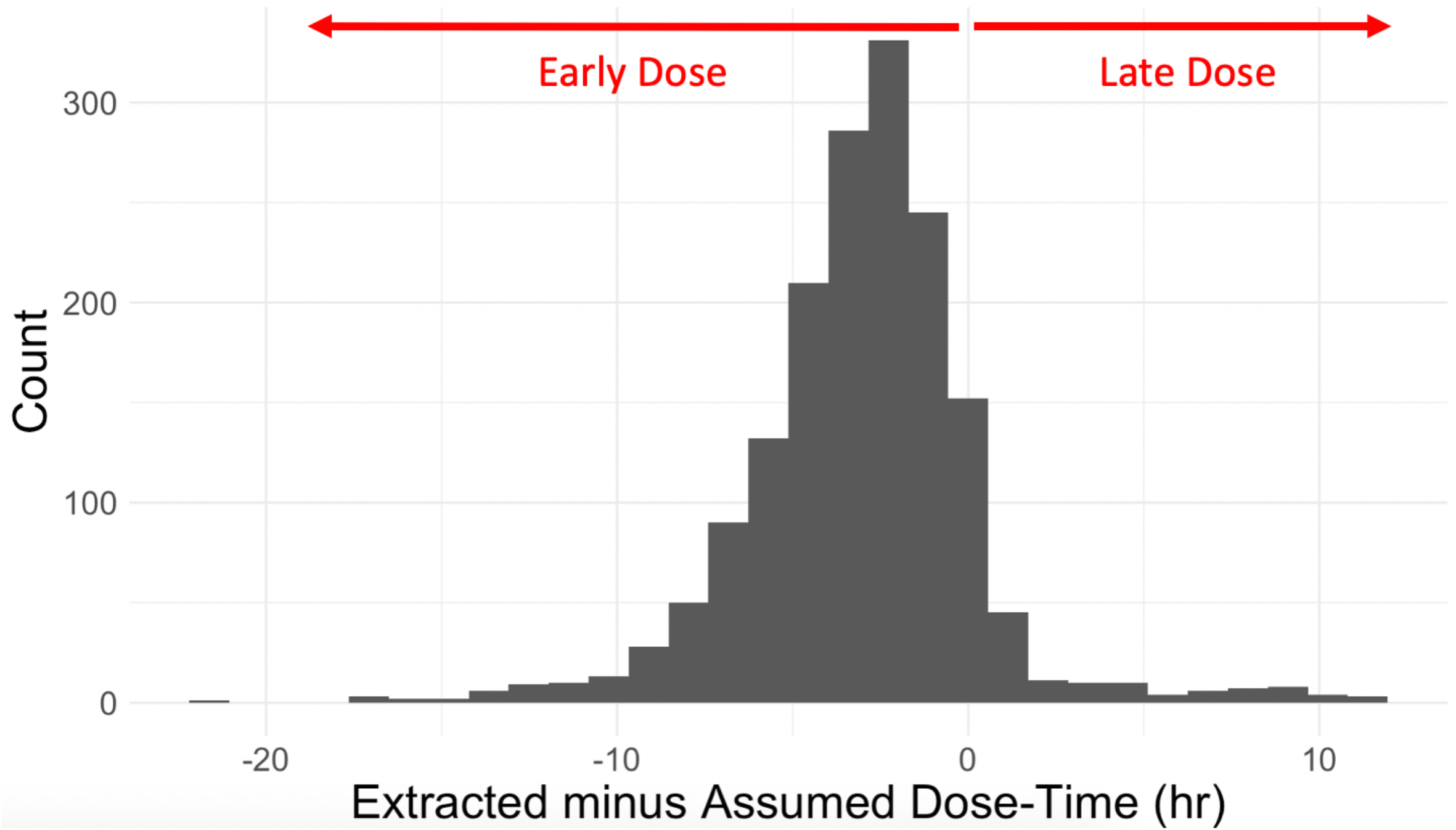
Differences in Extracted- and Assumed-Dose Time. Histogram depicting the distribution of the difference between extracted dose times and assumed dose times observed in the entire cohort. Negative values represent instances where the extracted dose time precedes the assumed dose time. Extracted dose times, which are likely close to actual dose times, tend to be earlier than assumed dose times; the median difference is 3 hours early. Extracted dose times which occur more than 12 hours before assumed dose times could potentially be due to a missed dose, likely accurately captured in the EHR.

### Sensitivity Analysis of k_a_ in Tacrolimus Population PK Model

**Table 4** shows parameter estimates from the entire cohort with last-dose time when *k*_*a*_ is either assumed to be 4.5, 3.09, or 0.375. The estimates (± SE) of typical values for *CL* at median covariate values and no *CYP3A5*1* allele were 26.2 ± 0.7, 26.2 ± 0.7, and 26.6 ± 0.7, and those for *V* were 3726 ± 844, 3730 ± 788, and 3633 ± 628 for *k*_*a*_ assumed to be 4.5, 3.09, and 0.375, respectively. None of the estimates for *k*_*a*_ = 3.09 and *k*_*a*_ = 0.375 models fall outside of the 95% Wald CIs for the corresponding parameters of the *k*_*a*_ = 4.5 model.

**Table 4.** Sensitivity Analysis. PK parameter estimates for models assuming *k*_*a*_ to be 4.5, 3.09, 0.375 (left to right). Estimates are presented as mean (SE) [Wald 95% confidence interval]. *SNP* is coded continuously as 1, 2, 3 for 0, 1, and 2 mutations, respectively. The *age*_*ij*_, *alb*_*ij*_, and *hgb*_*ij*_ represent covariates values for subject *i* at time *j. CL* and *V* are population PK parameters for clearance and volume of distribution in the base model. The *ω*_*CL*_ and *ω*_*V*_ are the between-subject variance components for clearance and volume of distribution, respectively, presented as %CV. The *σ*_*prop*_ and *σ*_*add*_ represent the proportional and additive residual errors in the combined residual error model, presented as %CV and the standard deviation, respectively. The *θ*s denote model parameters as typically used in statistical models.

| | $k_a = 4.5$ | $k_a = 3.09$ | $k_a = 0.375$ |
| --- | --- | --- | --- |
| Objective Function | 10169 | 10169 | 10170 |
| Parameter | Estimate (SE) [95% CI] |  |  |
| $CL_{ij} = \theta_1 \times SNP^{\theta_2} \times (age_{ij}/47)^{\theta_3} \times (alb_{ij}/4.1)^{\theta_4} \times (hgb_{ij}/12.5)^{\theta_5}$ | | | |
| $\theta_1$ | 26.2 (0.7)<br>[24.8, 27.6] | 26.2 (0.7)<br>[24.8, 27.6] | 26.6 (0.7)<br>[25.2, 28.1] |
| $\theta_2$ | 0.71 (0.05)<br>[0.61, 0.81] | 0.71 (0.05)<br>[0.61, 0.81] | 0.73 (0.05)<br>[0.63, 0.83] |
| $\theta_3$ | -0.26 (0.08)<br>[-0.42, -0.1] | -0.26 (0.08)<br>[-0.42, -0.10] | -0.26 (0.07)<br>[-0.40, -0.12] |
| $\theta_4$ | 0.35 (0.12)<br>[0.11, 0.59] | 0.35 (0.12)<br>[0.11, 0.59] | 0.36 (0.12)<br>[0.12, 0.60] |
| $\theta_5$ | -0.29 (0.09)<br>[-0.47, -0.11] | -0.29 (0.09)<br>[-0.47, -0.11] | -0.30 (0.09)<br>[-0.48, -0.12] |
| $V_i = \theta_6$ | | | |
| $\theta_6$ | 3726 (844)<br>[2072, 5379] | 3730 (788)<br>[2185, 5275] | 3633 (628)<br>[2401, 4864] |
| $\omega_{CL}$ (%CV) | 40.8 (1.9)<br>[37.1, 44.5] | 40.8 (1.8)<br>[37.2, 44.4] | 41.2 (1.6)<br>[38.0, 44.4] |
| $\omega_V$ (%CV) | 65.3 (22.2)<br>[21.7, 108.9] | 65.4 (21.4)<br>[23.6, 107.3] | 65.0 (6.4)<br>[52.5, 77.5] |
| $\sigma_{prop}$ (%CV) | 31.8 (1.2)<br>[29.4, 34.2] | 31.8 (1.2)<br>[29.4, 34.2] | 31.8 (1.2)<br>[29.4, 34.2] |
| $\sigma_{add}$ (ng/ml) | 0.52 (0.38)<br>[0, 1.26] | 0.52 (0.38)<br>[0, 1.26] | 0.52 (0.37)<br>[0, 1.25] |

### Simulation Study

**Table 5** and **Table 6** show the main results of the simulation study investigating the impact of misspecified last-dose times and incorrect specification of absorption rate constant *k*_*a*_, respectively, focusing on scenarios for high variability. We summarize our findings related to estimation of the main PK parameters, *CL, V*, and *k*_*e*_, but the results for variance components can also be found in the tables.

**Table 5.**
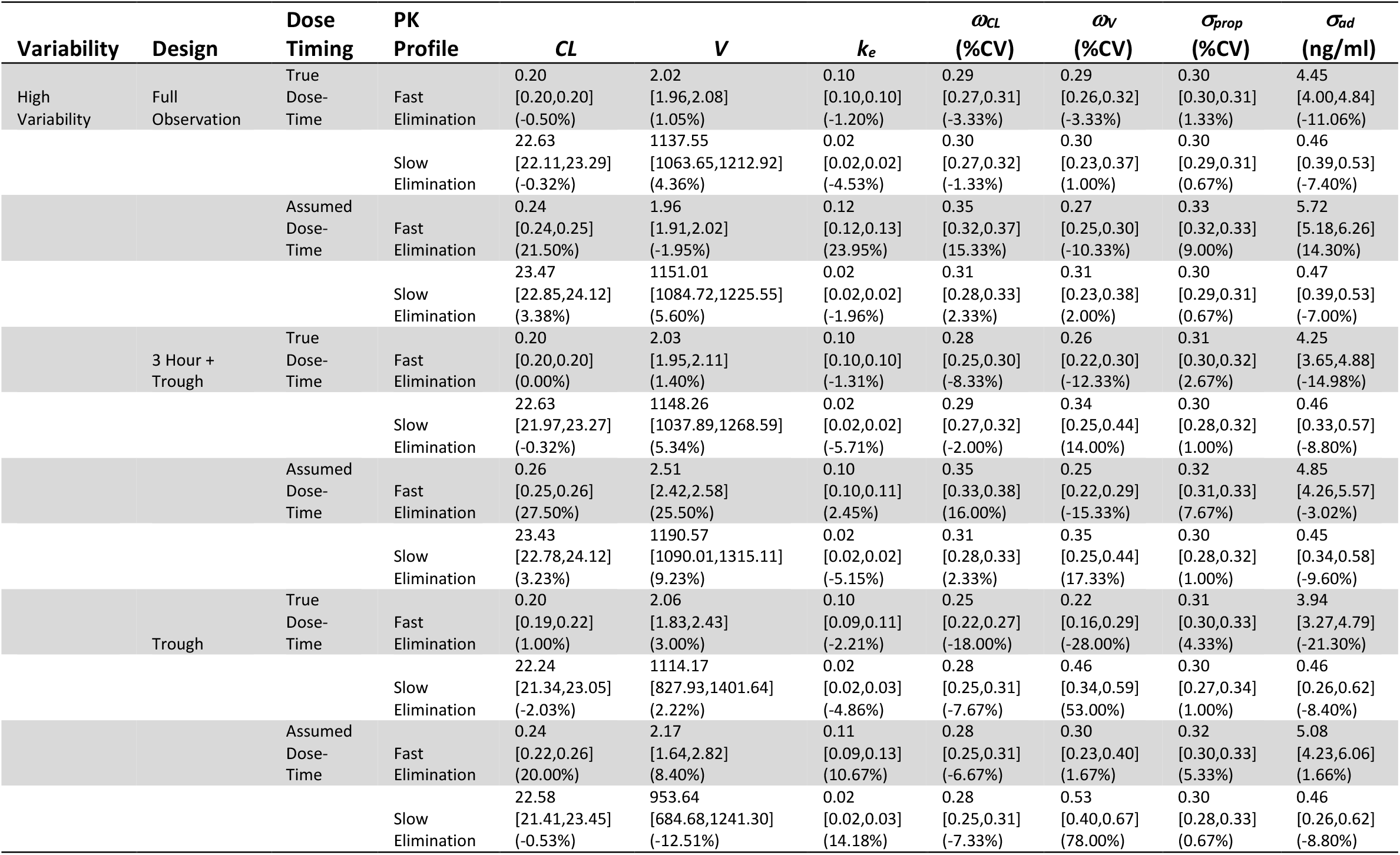
Simulation Results from 200 repetitions of a total of 12 scenarios: two PK profiles (SEL, FEL), one scenarios of variability (HV), three observation designs (FO, 3T, TR), and three assumed values for *k*_*a*_ (True *k*_*a*_, *k*_*a*_ x 10, *k*_*a*_ / 10). Simulation results are presented as medians, 10 to 90 percentiles in square brackets, and percent bias in parentheses. The *k*_*e*_, *CL*, and *V* represent population PK parameters for elimination rate constant, clearance, and volume of distribution respectively. The *ω*_*CL*_ and *ω*_*V*_ are the between-subject variance components for clearance and volume of distribution, presented as %CV. The *σ*_*prop*_ and *σ*_*add*_ represent the proportional and additive residual errors in the combined residual error model, presented as %CV and the standard deviation, respectively. True underlying parameters are shown in **Table 1**.

**Table 6.**
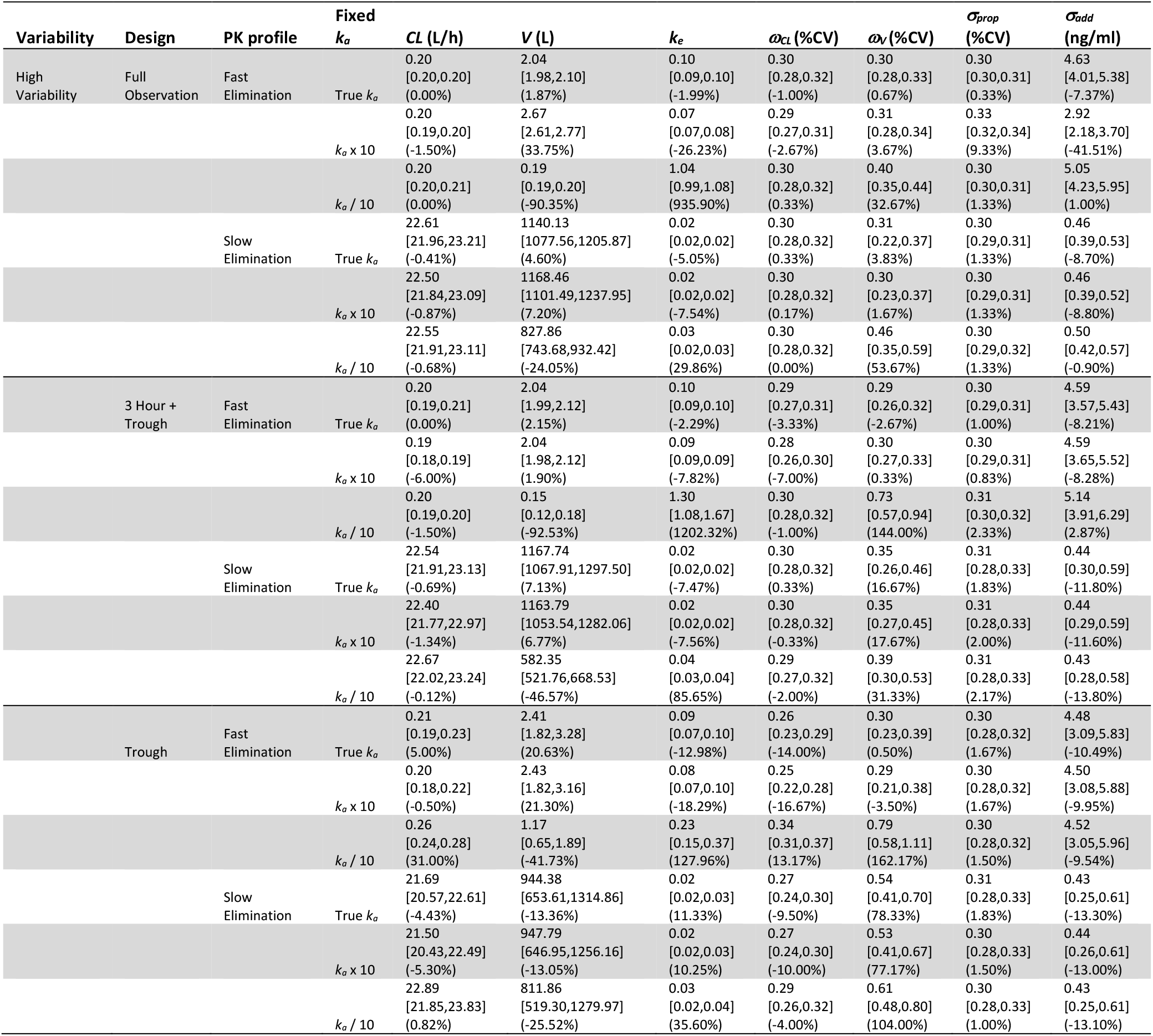
Simulation Results from 200 repetitions of a total of 18 scenarios: two PK profiles (SEL, FEL), one scenarios of variability (HV), three observation designs (FO, 3T, TR), and two last-dose times (AD, TD). Simulation results are presented as medians, 10 to 90 percentiles in square brackets, and percent bias in parentheses. The *k*_*e*_, *CL*, and *V* represent population PK parameters for elimination rate constant, clearance, and volume of distribution respectively. The *ω*_*CL*_ and *ω*_*V*_ are the between-subject variance components for clearance and volume of distribution, presented as %CV. The *σ*_*prop*_ and *σ*_*add*_ represent the proportional and additive residual errors in the combined residual error model, presented as %CV and the standard deviation, respectively. True underlying parameters are shown in **Table 1**.

#### Impact of misspecified last-dose times

In the FO design, estimates for *V* were stable in the presence of last-dose timing errors, and percent bias never exceeded 5.6% under any scenario. While the dose time errors had little impact on the estimate of *CL* and *k*_*e*_ for the SEL scenarios, they were overestimated more than 20% in the FEL. In the 3T design with the FEL scenarios, both *V* and *CL* were overestimated more than 20% in the presence of dose time errors, but the SEL scenarios resulted in only moderate bias (about 10%) for *V* and little bias for *CL* and *k*_*e*_.

Under the TR design, the dose time errors moderately impacted estimates of *V* and *k*_*e*_ in both the FEL and SEL scenarios. In the FEL scenarios, both were overestimated by about 10%, which would be the reason for magnified bias of 20% seen in estimates of *CL*. In the SEL scenarios, *V* was underestimated by ∼10% while *k*_*e*_ was overestimated ∼10%, but little bias in *CL* was observed.

#### Impact of incorrect specification of *k*_*a*_

In the FO design, the biases in estimates of *CL* were very small, less than 1.5% across all scenarios. On the other hand, *V* and *k*_*e*_ were highly sensitive to incorrect assumptions about *k*_*a*_; Estimates of *V* and *k*_*e*_ were substantially biased, with much larger bias under Low *k*_*a*_ than High *k*_*a*_, and this trend was magnified in the FEL compared to the SEL scenarios. In the 3T design, estimates for *CL* were slightly biased with maximum bias of -6% in the FEL with High *k*_*a*_ scenarios. Estimates for *V* and *k*_*e*_ were highly biased especially for Low *k*_*a*_ across all PK profiles (e.g., biases from -46% to -92% for *V*, from 86% to 1199% for *k*_*e*_). In the TR design, estimates of *CL* had little bias for most scenarios except for the case of FEL with High *k*_*a*_ (32% bias). However, *V* and *k*_*e*_ were substantially biased for all FEL scenarios, even when *k*_*a*_ was assumed correctly (18% bias for *V*). In the SEL scenarios, *V* and *k*_*e*_ was again substantially biased under Low *k*_*a*_, -25% and 36% bias for *V* and *k*_*e*_, respectively, but less biased under High *k*_*a*_.

We also studied the impact of misspecified last-dose times and incorrect specification of *k*_*a*_ on populations with low interindividual and residual variability, which were included as reference cases. Although dose time errors and incorrect specification of *k*_*a*_ still could yield bias in the low variability scenarios, in general their effects are much smaller compared to those with high variability except for in a few scenarios. The complete results including scenarios with low variability can be found in **Supplemental Material**.

## Discussion

Our tacrolimus population PK study performed with real-world data using solely EHR data reproduced the well-established relationship between the effect of rs776746 in *CYP3A5* and tacrolimus clearance. Although the typical value of *CL* is not completely comparable due to different covariate models used across studies, our study estimated *CL* of 26.2 [24.8-27.6] for a 47-year-old with no *\*1* allele in *CYP3A5*, albumin 4.1 g/dL, and hemoglobin 12.5 g/dL, while the other studies reported 15.9 [13.2–18.6]^9^ and 26.6 [18–35.2].^10^ The difference may be due to the difference in the study populations, especially different post-transplantation days. Our cohort included data from 30 days of post-transplantation up to 1,095 days (3 years) with median of 482 days, while the median (range) post-transplantation days were 14 (1–175) and 9 (0–95) from these two studies, which are much shorter than ours. In addition, the study population for Li *et al*.^9^ was liver transplant subjects, which would have additional impact on clearance as poor liver function would reduce tacrolimus clearance.^26,27^

The differential frequency distribution of *CYP3A5*1* genotype between populations of African ancestry (AA) and Northern European ancestry (NE) is well established and is apparent in our study population (**Figure 5**). It is well known that presence of *CYP3A5*1* increases clearance. We observed greater *CYP3A5*1* genotype prevalence in the AA population compared to the NE population. Thus, clearance for AA subjects is much higher than for NE subjects if *CYP3A5*1* genotype is not accounted for. However, once genotype is accounted for, the difference between PK parameters in NE and AA subjects is negligible; inclusion of additional race covariate did not sufficiently improve model fit to be included in our final covariate model. AA patients are known to require higher doses of tacrolimus, but it is unknown to what extent this is due to greater prevalence of *CYP3A5*1* rather than differences in bioavailability or absorption rates.^28^ Our results support that this difference is more attributable to the differential distribution of *CYP3A5* genotype than any other factors.

**Figure 5.**
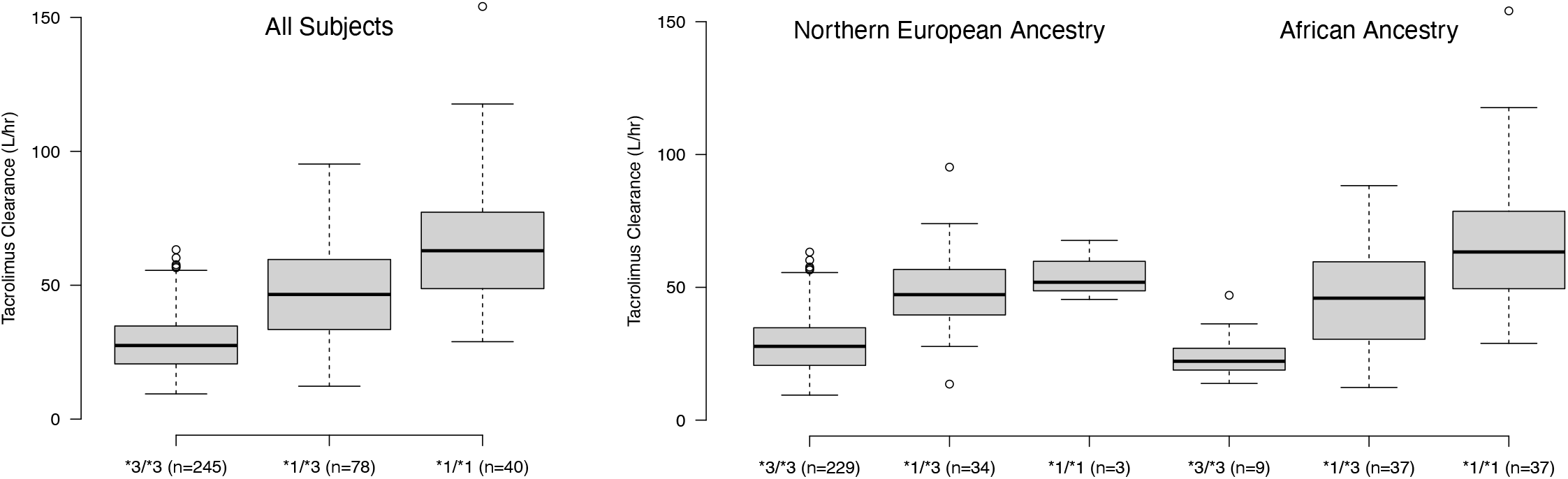
Estimated Clearance by *CYP3A5* Diplotype. Box and whisker plots for clearance by *CYP3A5* diplotype for all subjects (left) and clearance by *CYP3A5* diplotype among Northern European ancestry and African ancestry subjects (right).

The inclusion of extracted vs. assumed last-dosing time did not meaningfully alter the estimation of PK parameters or covariate effects in the analysis performed using datasets with either the entire cohort or the reduced cohort which oversampled subjects for which more last-dosing time information was available. There is more similarity within cohorts regardless of availability for the last-dose time data than between the entire and the reduced cohort given the last-dose time data status. The lack of a difference can be attributed to the following reasons. First, the estimated timing of the last dose may be close enough to the actual time – this is a reasonable assumption as the compliance of the organ transplant patient population is known to be high because of the serious consequence of organ rejection if they are not compliant to taking medication at scheduled intervals. Second, tacrolimus concentration levels are likely in the steady state after 1 month of organ transplantation, the time from which our data collection was started. In addition, as tacrolimus has a long half-life, trough concentrations would well approximate the average steady-state concentration,^29^ which would allow for the PK modeling to be less sensitive to deviation of actual dosing time. *V* is, however, consistently estimated to be larger in the without last-dose time datasets; perhaps this can be attributed to the increased uncertainty due to incorrect last-dose times. These results can inform the design of PK studies performed using real-world data source such as EHRs for medications having similar PK characteristics.^14^

Our findings support the general modeling approach used in several tacrolimus population PK studies,^9,10^ where *k*_*a*_ was fixed at a published value – our estimates for clearance and covariate effects were not sensitive to our selection for *k*_*a*_. The choice of *k*_*a*_ in a range of a 12-fold difference (from 0.375 to 4.5) had little impact on parameter estimates; all parameter estimates from the *k*_*a*_ = 3.09 or 0.375 models were within the 95% CIs of the corresponding parameters in the *k*_*a*_ = 4.5 model. This is likely due to the use of trough concentrations, which are taken well after the absorption phase is dominant and are therefore less impacted by the absorption process. Given the twice-daily dosing schedule and clinical goals of tacrolimus blood concentration maintenance via TDM, any reasonable selection for *k*_*a*_ is not likely to impact tacrolimus PK modeling. These *k*_*a*_ values were reported for immediate-release formulations for tacrolimus, which is the formulation used in our study. Notably, the models with *k*_*a*_ set to 4.5 and 3.09 are nearly identical while the model with *k*_*a*_ set to 0.375 yielded slight, but negligible differences. The extended-release formulation of tacrolimus is now on the market and our findings suggest that PK parameter estimation should not be highly sensitive to assumptions about absorption rate.

The simulation study gives some insight into the findings of our study. The effect of incorrectly assumed dose times is more impactful on PK parameter estimation in the FEL scenarios, especially when the experimental design only includes trough concentrations. In the SEL scenarios, the impact of last-dose time is smaller. Even with the TR design in the SEL profile (which is closest to our study design), we found bias of only about 10% when estimating *V* and *k*_*e*_, but *CL* is always well estimated. This suggests that, even when trough concentrations alone are collected, the population PK analysis for drugs with slower rates may provide a reasonable estimate for clearance. This finding should give confidence to clinical dosing decisions based on these estimates as clearance is most relevant parameter to determine proper dosage. On the other hand, estimates from the FEL scenarios are more sensitive to the dose time errors. Thus, the analysis for drugs with faster elimination than tacrolimus would warrant some caution as they could be more sensitive to dose-time errors than what we observed in our study.

Sensitivity to *k*_*a*_ was also greater in the FEL than in the SEL. We again attribute this to the faster elimination. In particular, the ratio of absorption to elimination rates, *k*_*a*_/*k*_*e*_, appears to determine the magnitude of bias. In the FEL scenarios, *k*_*a*_ is one order of magnitude greater than *k*_*e*_ so that when we assume *k*_*a*_ to be one tenth its true value, they become equal. We included this scenario as an extreme case. In the SEL scenarios where true *k*_*a*_ is two orders of magnitude greater, even if *k*_*a*_ is one tenth its true value, it is still much larger than *k*_*e*_ and hence the effect of *k*_*a*_ assumption is less profound. This is particularly true in the design which considers only trough concentrations. As long as *k*_*a*_ is sufficiently large relative to *k*_*e*_, estimation results would not be very sensitive to incorrect assumptions about *k*_*a*_. Another important factor determining the magnitude of bias due to the incorrect *k*_*a*_ assumptions is the sampling design. Sensitivity increases as we take more observations, such as in 3T or FO design, where observations are taken immediately following dosing, when absorption is still dominant. Fortunately, there is little need to fix *k*_*a*_ at a certain value (or assume *k*_*a*_ incorrectly) in the PK modeling when such data is available. In general, the scenarios of SEL were less sensitive to incorrect *k*_*a*_ assumptions than those of the FEL where estimates of *CL* could be biased up to 31%. The only significant bias in the SEL was the estimation of *V* and *k*_*e*_ when *k*_*a*_ was assumed to be lower than the true *k*_*a*_. Bias in *CL* never exceeded 5.3% in the SEL even when estimates of *V* and *k*_*e*_ were substantially biased. The reason for this is that the bias in the estimates of *V* and *k*_*e*_ are in opposite direction with similar magnitude and hence the biases seem to be canceled out, leading to little bias in CL. Again, all these results suggest that tacrolimus may be less sensitive to incorrect assumptions about *k*_*a*_ than a drug with faster elimination.

Our study has some limitations. First, the last-dose time information was not available for all drug concentration measurements. However, based on the results from the reduced cohort that included the last-dose time information for 73% of drug levels, it does not appear that including last-dose times in PK modeling will result in significant changes as long as medications share a similar PK profile with tacrolimus, such as a long half-life. Second, we did not specifically study medications with very different PK profiles compared to tacrolimus to investigate the effects of the last-dose time and the fixed *k*_*a*_ on PK parameter estimates. However, based on the simulation studies we performed, medications with a slow elimination rate (or a long half-life) would be less sensitive to dose time errors and incorrect assumptions about *k*_*a*_, and our results would provide some guidance on performing population PK analyses using real-world data such as EHRs.

Finally, this study used a mix of clinically validated data and data extracted by *medextractR*. We want to continue to move away from costly clinical validation and demonstrate that findings are reproducible with entirely automated EHR data extraction. For this reason, we will aim to replicate these results using our complete medication information pipeline and compare it to the results obtained through clinically validated data. Reproduction of PK parameters and covariate effects will indicate that our system is viable to replace costly manual information extraction to build PK data.

## Data Availability

Data is not publicly available.

## Funding

LC is supported by NIH/NIGMS (R01-GM124109).

## Conflict of Interest / Disclosure

The authors have no conflicts of interest to disclose.

## Author Contributions

All authors participated in critical review and revision of the final manuscript and approved the final manuscript draft.

MLW: Wrote manuscript, performed research, and analyzed data.

HLW: Performed research and contributed analytical tools.

CB: Contributed analytical tools.

LC: Designed research, wrote manuscript, performed research, and analyzed data.

## Notes

### Competing Interest Statement

The authors have declared no competing interest.

### Funding Statement

This work is supported by NIH/NIGMS (R01-GM124109).

### Author Declarations

This study was approved by the Vanderbilt Institutional Review Board.

