## Supplemental Tables for "Sensitivity of Estimated Tacrolimus Population Pharmacokinetic Profile to Inaccurate Assumptions about Dose Timing and Absorption: An Investigation in Real-World and Simulated Data"

**Table S1. Simulation Results from 200 repetitions of a total of 24 scenarios: two PK profiles (SEL, FEL), two scenarios of variability (LV, HV), three observation designs (FO, 3T, TR), and two last-dose times (AD, TD).** Simulation results are presented as medians, 10 to 90 percentiles in square brackets, and percent bias in parentheses. The $k_e$, $CL$, and $V$ represent population PK parameters for elimination rate constant, clearance, and volume of distribution respectively. The $\omega_{CL}$ and $\omega_V$ are the between-subject variance components for clearance and volume of distribution, presented as %CV. The $\sigma_{prop}$ and $\sigma_{add}$ represent the proportional and additive residual errors in the combined residual error model, presented as %CV and the standard deviation, respectively. True underlying parameters are shown in **Table 1**.

| Design | Dose Timing | PK Profile | Variability | $CL$ | $V$ | $k_e$ | $\omega_{CL}$ (%CV) | $\omega_V$ (%CV) | $\sigma_{prop}$ (%CV) | $\sigma_{ad}$ (ng/ml) |
|---|---|---|---|---|---|---|---|---|---|---|
| Full Observation | True Dose-Time | Fast Elimination | High Variability | 0.20<br>[0.20,0.20]<br>(-0.50%) | 2.02<br>[1.96,2.08]<br>(1.05%) | 0.10<br>[0.10,0.10]<br>(-1.20%) | 0.29<br>[0.27,0.31]<br>(-3.33%) | 0.29<br>[0.26,0.32]<br>(-3.33%) | 0.30<br>[0.30,0.31]<br>(1.33%) | 4.45<br>[4.00,4.84]<br>(-11.06%) |
| | | | Low Variability | 0.20<br>[0.20,0.20]<br>(0.00%) | 2.00<br>[1.99,2.01]<br>(0.00%) | 0.10<br>[0.10,0.10]<br>(0.05%) | 0.05<br>[0.04,0.05]<br>(-4.00%) | 0.05<br>[0.04,0.05]<br>(-6.00%) | 0.05<br>[0.05,0.05]<br>(0.00%) | 0.96<br>[0.86,1.11]<br>(-3.90%) |
| | | Slow Elimination | High Variability | 22.63<br>[22.11,23.29]<br>(-0.32%) | 1137.55<br>[1063.65,1212.92]<br>(4.36%) | 0.02<br>[0.02,0.02]<br>(-4.53%) | 0.30<br>[0.27,0.32]<br>(-1.33%) | 0.30<br>[0.23,0.37]<br>(1.00%) | 0.30<br>[0.29,0.31]<br>(0.67%) | 0.46<br>[0.39,0.53]<br>(-7.40%) |
| | | | Low Variability | 22.71<br>[22.61,22.81]<br>(0.03%) | 1092.09<br>[1079.00,1104.90]<br>(0.19%) | 0.02<br>[0.02,0.02]<br>(-0.07%) | 0.05<br>[0.04,0.05]<br>(-2.00%) | 0.06<br>[0.04,0.07]<br>(14.00%) | 0.05<br>[0.04,0.06]<br>(0.00%) | 0.10<br>[0.05,0.14]<br>(0.00%) |
| | Assumed Dose-Time | Fast Elimination | High Variability | 0.24<br>[0.24,0.25]<br>(21.50%) | 1.96<br>[1.91,2.02]<br>(-1.95%) | 0.12<br>[0.12,0.13]<br>(23.95%) | 0.35<br>[0.32,0.37]<br>(15.33%) | 0.27<br>[0.25,0.30]<br>(-10.33%) | 0.33<br>[0.32,0.33]<br>(9.00%) | 5.72<br>[5.18,6.26]<br>(14.30%) |
| | | | Low Variability | 0.24<br>[0.24,0.24]<br>(20.00%) | 1.92 [1.91,1.94] (-<br>3.80%) | 0.12<br>[0.12,0.13]<br>(24.61%) | 0.06<br>[0.06,0.06]<br>(18.00%) | 0.02<br>[0.01,0.02]<br>(-68.00%) | 0.16<br>[0.16,0.16]<br>(218.00%) | 0.00<br>[0.00,0.00]<br>(-100.00%) |
| | | Slow Elimination | High Variability | 23.47<br>[22.85,24.12]<br>(3.38%) | 1151.01<br>[1084.72,1225.55]<br>(5.60%) | 0.02<br>[0.02,0.02]<br>(-1.96%) | 0.31<br>[0.28,0.33]<br>(2.33%) | 0.31<br>[0.23,0.38]<br>(2.00%) | 0.30<br>[0.29,0.31]<br>(0.67%) | 0.47<br>[0.39,0.53]<br>(-7.00%) |
| | | | Low Variability | 23.46<br>[23.35,23.57]<br>(3.35%) | 1099.61<br>[1086.89,1114.54]<br>(0.88%) | 0.02<br>[0.02,0.02]<br>(2.47%) | 0.05<br>[0.05,0.05]<br>(2.00%) | 0.06<br>[0.04,0.07]<br>(16.00%) | 0.06<br>[0.05,0.06]<br>(14.00%) | 0.06<br>[0.01,0.10]<br>(-42.00%) |
| 3 Hour + Trough | True Dose-Time | Fast Elimination | High Variability | 0.20<br>[0.20,0.20]<br>(0.00%) | 2.03<br>[1.95,2.11]<br>(1.40%) | 0.10<br>[0.10,0.10]<br>(-1.31%) | 0.28<br>[0.25,0.30]<br>(-8.33%) | 0.26<br>[0.22,0.30]<br>(-12.33%) | 0.31<br>[0.30,0.32]<br>(2.67%) | 4.25<br>[3.65,4.88]<br>(-14.98%) |
| | | | Low Variability | 0.20<br>[0.20,0.20]<br>(0.00%) | 2.00<br>[1.99,2.01]<br>(-0.05%) | 0.10<br>[0.10,0.10]<br>(0.05%) | 0.04<br>[0.04,0.05]<br>(-10.00%) | 0.04<br>[0.04,0.05]<br>(-16.00%) | 0.05<br>[0.05,0.05]<br>(2.00%) | 0.95<br>[0.76,1.12]<br>(-4.60%) |
| | | Slow Elimination | High Variability | 22.63<br>[21.97,23.27]<br>(-0.32%) | 1148.26<br>[1037.89,1268.59]<br>(5.34%) | 0.02<br>[0.02,0.02]<br>(-5.71%) | 0.29<br>[0.27,0.32]<br>(-2.00%) | 0.34<br>[0.25,0.44]<br>(14.00%) | 0.30<br>[0.28,0.32]<br>(1.00%) | 0.46<br>[0.33,0.57]<br>(-8.80%) |
| | | | Low Variability | 22.70<br>[22.60,22.82]<br>(0.00%) | 1091.94<br>[1073.77,1114.99]<br>(0.18%) | 0.02<br>[0.02,0.02]<br>(-0.29%) | 0.05<br>[0.04,0.05]<br>(-2.00%) | 0.07<br>[0.05,0.09]<br>(38.00%) | 0.05<br>[0.04,0.06]<br>(4.00%) | 0.08<br>[0.02,0.16]<br>(-16.00%) |
| | Assumed Dose-Time | Fast Elimination | High Variability | 0.26<br>[0.25,0.26]<br>(27.50%) | 2.51<br>[2.42,2.58]<br>(25.50%) | 0.10<br>[0.10,0.11]<br>(2.45%) | 0.35<br>[0.33,0.38]<br>(16.00%) | 0.25<br>[0.22,0.29]<br>(-15.33%) | 0.32<br>[0.31,0.33]<br>(7.67%) | 4.85<br>[4.26,5.57]<br>(-3.02%) |
| | | | Low Variability | 0.25<br>[0.25,0.25]<br>(26.00%) | 2.40 [2.38,2.42]<br>(19.95%) | 0.11<br>[0.10,0.11]<br>(5.11%) | 0.06<br>[0.06,0.06]<br>(20.00%) | 0.02<br>[0.01,0.02]<br>(-68.00%) | 0.11<br>[0.11,0.12]<br>(128.00%) | 1.54<br>[1.22,1.86]<br>(53.80%) |
| | | Slow Elimination | High Variability | 23.43<br>[22.78,24.12]<br>(3.23%) | 1190.57<br>[1090.01,1315.11]<br>(9.23%) | 0.02<br>[0.02,0.02]<br>(-5.15%) | 0.31<br>[0.28,0.33]<br>(2.33%) | 0.35<br>[0.25,0.44]<br>(17.33%) | 0.30<br>[0.28,0.32]<br>(1.00%) | 0.45<br>[0.34,0.58]<br>(-9.60%) |
| | | | Low Variability | 23.49<br>[23.38,23.60]<br>(3.47%) | 1130.15<br>[1112.99,1154.41]<br>(3.68%) | 0.02<br>[0.02,0.02]<br>(-0.27%) | 0.05<br>[0.05,0.05]<br>(2.00%) | 0.07<br>[0.05,0.09]<br>(42.00%) | 0.05<br>[0.04,0.06]<br>(8.00%) | 0.08<br>[0.02,0.16]<br>(-18.00%) |
| Trough | True Dose-Time | Fast Elimination | High Variability | 0.20<br>[0.19,0.22]<br>(1.00%) | 2.06<br>[1.83,2.43]<br>(3.00%) | 0.10<br>[0.09,0.11]<br>(-2.21%) | 0.25<br>[0.22,0.27]<br>(-18.00%) | 0.22<br>[0.16,0.29]<br>(-28.00%) | 0.31<br>[0.30,0.33]<br>(4.33%) | 3.94<br>[3.27,4.79]<br>(-21.30%) |
| | | | Low Variability | 0.20<br>[0.20,0.20]<br>(0.00%) | 2.00<br>[1.95,2.05]<br>(-0.15%) | 0.10<br>[0.10,0.10]<br>(0.00%) | 0.04<br>[0.03,0.04]<br>(-22.00%) | 0.04<br>[0.03,0.05]<br>(-18.00%) | 0.05<br>[0.04,0.06]<br>(0.00%) | 1.00<br>[0.31,1.46]<br>(-0.50%) |
| | | Slow Elimination | High Variability | 22.24<br>[21.34,23.05]<br>(-2.03%) | 1114.17<br>[827.93,1401.64]<br>(2.22%) | 0.02<br>[0.02,0.03]<br>(-4.86%) | 0.28<br>[0.25,0.31]<br>(-7.67%) | 0.46<br>[0.34,0.59]<br>(53.00%) | 0.30<br>[0.27,0.34]<br>(1.00%) | 0.46<br>[0.26,0.62]<br>(-8.40%) |
| | | | Low Variability | 22.68<br>[22.44,23.01]<br>(-0.09%) | 1103.43<br>[1004.29,1233.25]<br>(1.23%) | 0.02<br>[0.02,0.02]<br>(-1.14%) | 0.04<br>[0.04,0.05]<br>(-14.00%) | 0.17<br>[0.12,0.20]<br>(232.00%) | 0.05<br>[0.02,0.06]<br>(2.00%) | 0.09<br>[0.00,0.26]<br>(-7.00%) |
| | Assumed Dose-Time | Fast Elimination | High Variability | 0.24<br>[0.22,0.26]<br>(20.00%) | 2.17<br>[1.64,2.82]<br>(8.40%) | 0.11<br>[0.09,0.13]<br>(10.67%) | 0.28<br>[0.25,0.31]<br>(-6.67%) | 0.30<br>[0.23,0.40]<br>(1.67%) | 0.32<br>[0.30,0.33]<br>(5.33%) | 5.08<br>[4.23,6.06]<br>(1.66%) |
| | | | Low Variability | 0.21<br>[0.20,0.22]<br>(4.00%) | 1.48<br>[1.32,1.61]<br>(-26.10%) | 0.14<br>[0.13,0.15]<br>(41.10%) | 0.03<br>[0.02,0.04]<br>(-34.00%) | 0.07<br>[0.05,0.09]<br>(48.00%) | 0.13<br>[0.09,0.14]<br>(162.00%) | 0.60<br>[0.01,3.29]<br>(-39.50%) |
| | | Slow Elimination | High Variability | 22.58<br>[21.41,23.45]<br>(-0.53%) | 953.64<br>[684.68,1241.30]<br>(-12.51%) | 0.02<br>[0.02,0.03]<br>(14.18%) | 0.28<br>[0.25,0.31]<br>(-7.33%) | 0.53<br>[0.40,0.67]<br>(78.00%) | 0.30<br>[0.28,0.33]<br>(0.67%) | 0.46<br>[0.26,0.62]<br>(-8.80%) |
| | | | Low Variability | 23.26<br>[22.91,23.61]<br>(2.47%) | 1066.20<br>[915.55,1220.93]<br>(-2.18%) | 0.02<br>[0.02,0.03]<br>(4.82%) | 0.04<br>[0.04,0.05]<br>(-12.00%) | 0.22<br>[0.16,0.28]<br>(342.00%) | 0.05<br>[0.02,0.06]<br>(0.00%) | 0.10<br>[0.00,0.28]<br>(1.00%) |

**Table S2. Simulation Results from 200 repetitions of a total of 36 scenarios: two PK profiles (SEL, FEL), two scenarios of variability (LV, HV), three observation designs (FO, 3T, TR), and three assumed values for $k_a$ (True $k_a$, $k_a$ x 10, $k_a$ / 10).** Simulation results are presented as medians, 10 to 90 percentiles in square brackets, and percent bias in parentheses. The $k_e$, $CL$, and $V$ represent population PK parameters for elimination rate constant, clearance, and volume of distribution respectively. The $\omega_{CL}$ and $\omega_V$ are the between-subject variance components for clearance and volume of distribution, presented as %CV. The $\sigma_{prop}$ and $\sigma_{add}$ represent the proportional and additive residual errors in the combined residual error model, presented as %CV and the standard deviation, respectively. True underlying parameters are shown in **Table 1**.

| Design | PK profile | Variability | Fixed $k_a$ | $CL$ (L/h) | $V$ (L) | $k_e$ | $\omega_{CL}$ (%CV) | $\omega_V$ (%CV) | $\sigma_{prop}$ (%CV) | $\sigma_{add}$ (ng/ml) |
|---|---|---|---|---|---|---|---|---|---|---|
| Full Observation | Fast Elimination | High Variability | True $k_a$ | 0.20 [0.20,0.20] (0.00%) | 2.04 [1.98,2.10] (1.87%) | 0.10 [0.09,0.10] (-1.99%) | 0.30 [0.28,0.32] (-1.00%) | 0.30 [0.28,0.33] (0.67%) | 0.30 [0.30,0.31] (0.33%) | 4.63 [4.01,5.38] (-7.37%) |
| | | | $k_a$ x 10 | 0.20 [0.19,0.20] (-1.50%) | 2.67 [2.61,2.77] (33.75%) | 0.07 [0.07,0.08] (-26.23%) | 0.29 [0.27,0.31] (-2.67%) | 0.31 [0.28,0.34] (3.67%) | 0.33 [0.32,0.34] (9.33%) | 2.92 [2.18,3.70] (-41.51%) |
| | | | $k_a$ / 10 | 0.20 [0.20,0.21] (0.00%) | 0.19 [0.19,0.20] (-90.35%) | 1.04 [0.99,1.08] (935.90%) | 0.30 [0.28,0.32] (0.33%) | 0.40 [0.35,0.44] (32.67%) | 0.30 [0.30,0.31] (1.33%) | 5.05 [4.23,5.95] (1.00%) |
| | | Low Variability | True $k_a$ | 0.20 [0.20,0.20] (0.00%) | 2.00 [1.99,2.01] (0.05%) | 0.10 [0.10,0.10] (-0.10%) | 0.05 [0.05,0.05] (-2.00%) | 0.05 [0.04,0.05] (-2.00%) | 0.05 [0.05,0.05] (0.00%) | 0.99 [0.77,1.18] (-1.10%) |
| | | | $k_a$ x 10 | 0.20 [0.20,0.20] (-1.00%) | 2.59 [2.57,2.60] (29.40%) | 0.08 [0.08,0.08] (-23.66%) | 0.05 [0.04,0.05] (-8.00%) | 0.02 [0.02,0.03] (-54.00%) | 0.10 [0.10,0.10] (102.00%) | 0.00 [0.00,0.00] (-100.00%) |
| | | | $k_a$ / 10 | 0.20 [0.20,0.20] (0.00%) | 0.20 [0.20,0.20] (-90.00%) | 1.00 [1.00,1.01] (900.00%) | 0.05 [0.05,0.05] (0.00%) | 0.06 [0.05,0.07] (18.00%) | 0.05 [0.05,0.05] (-2.00%) | 1.22 [1.02,1.43] (22.35%) |
| | Slow Elimination | High Variability | True $k_a$ | 22.61 [21.96,23.21] (-0.41%) | 1140.13 [1077.56,1205.87] (4.60%) | 0.02 [0.02,0.02] (-5.05%) | 0.30 [0.28,0.32] (0.33%) | 0.31 [0.22,0.37] (3.83%) | 0.30 [0.29,0.31] (1.33%) | 0.46 [0.39,0.53] (-8.70%) |
| | | | $k_a$ x 10 | 22.50 [21.84,23.09] (-0.87%) | 1168.46 [1101.49,1237.95] (7.20%) | 0.02 [0.02,0.02] (-7.54%) | 0.30 [0.28,0.32] (0.17%) | 0.30 [0.23,0.37] (1.67%) | 0.30 [0.29,0.31] (1.33%) | 0.46 [0.39,0.52] (-8.80%) |
| | | | $k_a$ / 10 | 22.55 [21.91,23.11] (-0.68%) | 827.86 [743.68,932.42] (-24.05%) | 0.03 [0.02,0.03] (29.86%) | 0.30 [0.28,0.32] (0.00%) | 0.46 [0.35,0.59] (53.67%) | 0.30 [0.29,0.32] (1.33%) | 0.50 [0.42,0.57] (-0.90%) |
| | | Low Variability | True $k_a$ | 22.70 [22.59,22.79] (0.00%) | 1092.74 [1081.33,1104.30] (0.25%) | 0.02 [0.02,0.02] (-0.27%) | 0.05 [0.05,0.05] (0.00%) | 0.06 [0.04,0.08] (20.00%) | 0.05 [0.04,0.06] (1.00%) | 0.09 [0.05,0.14] (-6.00%) |
| | | | $k_a$ x 10 | 22.60 [22.49,22.69] (-0.44%) | 1128.54 [1115.35,1139.60] (3.54%) | 0.02 [0.02,0.02] (-3.77%) | 0.05 [0.05,0.05] (0.00%) | 0.06 [0.04,0.07] (16.00%) | 0.05 [0.04,0.06] (2.00%) | 0.09 [0.05,0.14] (-10.50%) |
| | | | $k_a$ / 10 | 22.65 [22.54,22.75] (-0.22%) | 760.05 [744.25,784.20] (-30.27%) | 0.03 [0.03,0.03] (43.05%) | 0.05 [0.05,0.05] (-2.00%) | 0.07 [0.05,0.08] (32.00%) | 0.00 [0.00,0.00] (-100.00%) | 0.64 [0.63,0.64] (536.00%) |

| Design | PK profile | Variability | Fixed $k_a$ | $CL$ (L/h) | $V$ (L) | $k_e$ | $\omega_{CL}$ (%CV) | $\omega_V$ (%CV) | $\sigma_{prop}$ (%CV) | $\sigma_{add}$ (ng/ml) |
|---|---|---|---|---|---|---|---|---|---|---|
| 3 Hour + Trough | Fast Elimination | High Variability | True $k_a$ | 0.20<br>[0.19,0.21]<br>(0.00%) | 2.04<br>[1.99,2.12]<br>(2.15%) | 0.10<br>[0.09,0.10]<br>(-2.29%) | 0.29<br>[0.27,0.31]<br>(-3.33%) | 0.29<br>[0.26,0.32]<br>(-2.67%) | 0.30<br>[0.29,0.31]<br>(1.00%) | 4.59<br>[3.57,5.43]<br>(-8.21%) |
| | | | $k_a$ x 10 | 0.19<br>[0.18,0.19]<br>(-6.00%) | 2.04<br>[1.98,2.12]<br>(1.90%) | 0.09<br>[0.09,0.09]<br>(-7.82%) | 0.28<br>[0.26,0.30]<br>(-7.00%) | 0.30<br>[0.27,0.33]<br>(0.33%) | 0.30<br>[0.29,0.31]<br>(0.83%) | 4.59<br>[3.65,5.52]<br>(-8.28%) |
| | | | $k_a$ / 10 | 0.20<br>[0.19,0.20]<br>(-1.50%) | 0.15<br>[0.12,0.18]<br>(-92.53%) | 1.30<br>[1.08,1.67]<br>(1202.32%) | 0.30<br>[0.28,0.32]<br>(-1.00%) | 0.73<br>[0.57,0.94]<br>(144.00%) | 0.31<br>[0.30,0.32]<br>(2.33%) | 5.14<br>[3.91,6.29]<br>(2.87%) |
| | | Low Variability | True $k_a$ | 0.20<br>[0.20,0.20]<br>(0.00%) | 2.00<br>[1.99,2.01]<br>(0.05%) | 0.10<br>[0.10,0.10]<br>(-0.10%) | 0.05<br>[0.04,0.05]<br>(-4.00%) | 0.05<br>[0.04,0.05]<br>(-6.00%) | 0.05<br>[0.05,0.05]<br>(2.00%) | 0.94<br>[0.68,1.18]<br>(-6.20%) |
| | | | $k_a$ x 10 | 0.19<br>[0.19,0.19]<br>(-5.50%) | 2.01<br>[2.00,2.02]<br>(0.30%) | 0.09<br>[0.09,0.09]<br>(-6.06%) | 0.05<br>[0.04,0.05]<br>(-8.00%) | 0.05<br>[0.04,0.05]<br>(-4.00%) | 0.05<br>[0.05,0.05]<br>(2.00%) | 0.94<br>[0.68,1.17]<br>(-6.50%) |
| | | | $k_a$ / 10 | 0.20<br>[0.20,0.20]<br>(-0.50%) | 0.19<br>[0.18,0.19]<br>(-90.60%) | 1.06<br>[1.03,1.09]<br>(958.51%) | 0.05<br>[0.05,0.05]<br>(-2.00%) | 0.24<br>[0.20,0.26]<br>(373.00%) | 0.05<br>[0.05,0.05]<br>(0.00%) | 1.07<br>[0.84,1.35]<br>(7.50%) |
| | Slow Elimination | High Variability | True $k_a$ | 22.54<br>[21.91,23.13]<br>(-0.69%) | 1167.74<br>[1067.91,1297.50]<br>(7.13%) | 0.02<br>[0.02,0.02]<br>(-7.47%) | 0.30<br>[0.28,0.32]<br>(0.33%) | 0.35<br>[0.26,0.46]<br>(16.67%) | 0.31<br>[0.28,0.33]<br>(1.83%) | 0.44<br>[0.30,0.59]<br>(-11.80%) |
| | | | $k_a$ x 10 | 22.40<br>[21.77,22.97]<br>(-1.34%) | 1163.79<br>[1053.54,1282.06]<br>(6.77%) | 0.02<br>[0.02,0.02]<br>(-7.56%) | 0.30<br>[0.28,0.32]<br>(-0.33%) | 0.35<br>[0.27,0.45]<br>(17.67%) | 0.31<br>[0.28,0.33]<br>(2.00%) | 0.44<br>[0.29,0.59]<br>(-11.60%) |
| | | | $k_a$ / 10 | 22.67<br>[22.02,23.24]<br>(-0.12%) | 582.35<br>[521.76,668.53]<br>(-46.57%) | 0.04<br>[0.03,0.04]<br>(85.65%) | 0.29<br>[0.27,0.32]<br>(-2.00%) | 0.39<br>[0.30,0.53]<br>(31.33%) | 0.31<br>[0.28,0.33]<br>(2.17%) | 0.43<br>[0.28,0.58]<br>(-13.80%) |
| | | Low Variability | True $k_a$ | 22.69<br>[22.59,22.79]<br>(-0.03%) | 1094.52<br>[1075.98,1113.57]<br>(0.41%) | 0.02<br>[0.02,0.02]<br>(-0.53%) | 0.05<br>[0.05,0.05]<br>(0.00%) | 0.07<br>[0.05,0.10]<br>(46.00%) | 0.05<br>[0.04,0.06]<br>(2.00%) | 0.09<br>[0.02,0.16]<br>(-7.00%) |
| | | | $k_a$ x 10 | 22.55<br>[22.45,22.65]<br>(-0.65%) | 1088.20<br>[1069.74,1108.19]<br>(-0.17%) | 0.02<br>[0.02,0.02]<br>(-0.52%) | 0.05<br>[0.05,0.05]<br>(-2.00%) | 0.07<br>[0.06,0.10]<br>(44.00%) | 0.05<br>[0.04,0.06]<br>(2.00%) | 0.09<br>[0.02,0.16]<br>(-7.50%) |
| | | | $k_a$ / 10 | 22.67<br>[22.56,22.76]<br>(-0.15%) | 510.93<br>[503.33,520.70]<br>(-53.13%) | 0.04<br>[0.04,0.05]<br>(112.73%) | 0.05<br>[0.04,0.05]<br>(-2.00%) | 0.07<br>[0.05,0.09]<br>(40.00%) | 0.05<br>[0.04,0.06]<br>(2.00%) | 0.09<br>[0.02,0.16]<br>(-9.00%) |

| Design | PK profile | Variability | Fixed $k_a$ | *CL* (L/h) | *V* (L) | $k_e$ | $\omega_{CL}$ (%CV) | $\omega_V$ (%CV) | $\sigma_{prop}$ (%CV) | $\sigma_{add}$ (ng/ml) |
|---|---|---|---|---|---|---|---|---|---|---|
| Trough | Fast Elimination | High Variability | True $k_a$ | 0.21<br>[0.19,0.23]<br>(5.00%) | 2.41<br>[1.82,3.28]<br>(20.63%) | 0.09<br>[0.07,0.10]<br>(-12.98%) | 0.26<br>[0.23,0.29]<br>(-14.00%) | 0.30<br>[0.23,0.39]<br>(0.50%) | 0.30<br>[0.28,0.32]<br>(1.67%) | 4.48<br>[3.09,5.83]<br>(-10.49%) |
| | | | $k_a$ x 10 | 0.20<br>[0.18,0.22]<br>(-0.50%) | 2.43<br>[1.82,3.16]<br>(21.30%) | 0.08<br>[0.07,0.10]<br>(-18.29%) | 0.25<br>[0.22,0.28]<br>(-16.67%) | 0.29<br>[0.21,0.38]<br>(-3.50%) | 0.30<br>[0.28,0.32]<br>(1.67%) | 4.50<br>[3.08,5.88]<br>(-9.95%) |
| | | | $k_a$ / 10 | 0.26<br>[0.24,0.28]<br>(31.00%) | 1.17<br>[0.65,1.89]<br>(-41.73%) | 0.23<br>[0.15,0.37]<br>(127.96%) | 0.34<br>[0.31,0.37]<br>(13.17%) | 0.79<br>[0.58,1.11]<br>(162.17%) | 0.30<br>[0.28,0.32]<br>(1.50%) | 4.52<br>[3.05,5.96]<br>(-9.54%) |
| | | Low Variability | True $k_a$ | 0.18<br>[0.18,0.19]<br>(-9.00%) | 1.54<br>[1.40,1.70]<br>(-22.75%) | 0.12<br>[0.11,0.13]<br>(18.14%) | 0.03<br>[0.02,0.04]<br>(-36.00%) | 0.07<br>[0.05,0.09]<br>(34.00%) | 0.05<br>[0.03,0.06]<br>(2.00%) | 0.93<br>[0.02,2.89]<br>(-7.05%) |
| | | | $k_a$ x 10 | 0.17<br>[0.16,0.18]<br>(-14.50%) | 1.55<br>[1.43,1.71]<br>(-22.38%) | 0.11<br>[0.10,0.12]<br>(9.93%) | 0.03<br>[0.02,0.04]<br>(-38.00%) | 0.06<br>[0.04,0.08]<br>(22.00%) | 0.05<br>[0.03,0.06]<br>(2.00%) | 0.93<br>[0.02,2.95]<br>(-6.80%) |
| | | | $k_a$ / 10 | 0.27<br>[0.26,0.28]<br>(35.00%) | 1.44<br>[1.17,1.87]<br>(-28.00%) | 0.19<br>[0.15,0.22]<br>(88.07%) | 0.05<br>[0.04,0.05]<br>(-4.00%) | 0.24<br>[0.17,0.32]<br>(382.00%) | 0.05<br>[0.03,0.06]<br>(2.00%) | 0.94<br>[0.03,2.91]<br>(-6.35%) |
| | Slow Elimination | High Variability | True $k_a$ | 21.69<br>[20.57,22.61]<br>(-4.43%) | 944.38<br>[653.61,1314.86]<br>(-13.36%) | 0.02<br>[0.02,0.03]<br>(11.33%) | 0.27<br>[0.24,0.30]<br>(-9.50%) | 0.54<br>[0.41,0.70]<br>(78.33%) | 0.31<br>[0.28,0.33]<br>(1.83%) | 0.43<br>[0.25,0.61]<br>(-13.30%) |
| | | | $k_a$ x 10 | 21.50<br>[20.43,22.49]<br>(-5.30%) | 947.79<br>[646.95,1256.16]<br>(-13.05%) | 0.02<br>[0.02,0.03]<br>(10.25%) | 0.27<br>[0.24,0.30]<br>(-10.00%) | 0.53<br>[0.41,0.67]<br>(77.17%) | 0.30<br>[0.28,0.33]<br>(1.50%) | 0.44<br>[0.26,0.61]<br>(-13.00%) |
| | | | $k_a$ / 10 | 22.89<br>[21.85,23.83]<br>(0.82%) | 811.86<br>[519.30,1279.97]<br>(-25.52%) | 0.03<br>[0.02,0.04]<br>(35.60%) | 0.29<br>[0.26,0.32]<br>(-4.00%) | 0.61<br>[0.48,0.80]<br>(104.00%) | 0.30<br>[0.28,0.33]<br>(1.00%) | 0.43<br>[0.25,0.61]<br>(-13.10%) |
| | | Low Variability | True $k_a$ | 22.54<br>[22.12,22.82]<br>(-0.69%) | 1052.08<br>[883.92,1190.57]<br>(-3.48%) | 0.02<br>[0.02,0.03]<br>(2.87%) | 0.04<br>[0.04,0.05]<br>(-15.00%) | 0.21<br>[0.16,0.27]<br>(327.00%) | 0.05<br>[0.02,0.06]<br>(2.00%) | 0.09<br>[0.00,0.29]<br>(-6.00%) |
| | | | $k_a$ x 10 | 22.43<br>[22.01,22.69]<br>(-1.17%) | 1059.12<br>[896.54,1180.71]<br>(-2.83%) | 0.02<br>[0.02,0.02]<br>(1.76%) | 0.04<br>[0.04,0.05]<br>(-14.00%) | 0.21<br>[0.16,0.26]<br>(312.00%) | 0.05<br>[0.02,0.06]<br>(2.00%) | 0.09<br>[0.00,0.29]<br>(-6.00%) |
| | | | $k_a$ / 10 | 23.67<br>[23.39,23.89]<br>(4.26%) | 975.26<br>[811.05,1145.94]<br>(-10.53%) | 0.02<br>[0.02,0.03]<br>(16.48%) | 0.05<br>[0.04,0.05]<br>(-6.00%) | 0.31<br>[0.23,0.40]<br>(519.00%) | 0.05<br>[0.02,0.06]<br>(1.00%) | 0.10<br>[0.00,0.29]<br>(-0.50%) |
